# Patterns of Muscle Health in Single- and Multi-Site Chronic Pain: A UK Biobank Normative Modeling Study

**DOI:** 10.64898/2026.06.19.26356062

**Authors:** Merve Kaptan, Yiyu Wang, Augustijn de Boer, Ananya Goyal, Skylar Holmes, Kerem Ozkan, Sandrine Bédard, Teresa Indriolo, Christine S W Law, Dario Pfyffer, Joel Fundaun, Estifanos Berhe, Garry E. Gold, Akshay Chaudhari, Pai S Anoosha, Anthony A. Gatti, Feliks Kogan, Brian A. Hargreaves, Scott L. Delp, John Ratliff, Serena Hu, Anand Veeravagu, Atman Desai, Suzanne Tharin, Todd Alamin, Andrew C. Smith, Marnee J. McKay, Brian Kim, Robert Walsh, Alec Schielke, Dean Dennis, Johannes Decker, Benjamin De Leener, Julien Cohen-Adad, Zachary A. Smith, Fauziyya Muhammad, James M. Elliott, Andre F. Marquand, Sean Mackey, Evert Onno Wesselink, Kenneth A. Weber

## Abstract

**Background:** Chronic pain is associated with impaired muscle health, but whether these changes reflect site-specific factors, broader systemic factors, or both remains unclear. The purpose of this study is to determine whether normative markers of muscle health derived from MRI show site-specific patterns in chronic pain.

**Methods:** UK Biobank participants who underwent whole-body MRI from 2006 to 2010 were included in this retrospective cross-sectional study. The MuscleMap Toolbox quantified volume and intramuscular fat (IMF) in 42 muscles of the abdomen, pelvis, and thigh. Normative models trained on a no pain group generated muscle-specific deviations from normal (i.e., Z-scores) for single- and multi-site chronic and acute pain.

**Results:** Of 17,843 participants, the primary site-specific analysis included 9,704 no pain, 885 single-site chronic back pain (CBP), 438 single-site chronic hip pain (CHP), and 1,315 single-site chronic knee pain (CKP) participants (n=12,342; mean age 63.7±7.5 years; 52.7% female). Additional analyses included single-site chronic neck/shoulder pain, acute pain, and multi-site chronic pain groups. In CBP, deviations were localized to abdominal muscles, with decreased volume in 6/8 and increased IMF in 6/8. In CHP, deviations were broad, with decreased volume in 3/8 of the abdominal and 14/26 of the thigh muscles, and increased IMF in 6/8 of the abdominal, 5/8 of the pelvic, and 4/26 of the thigh muscles. In CKP, deviations were localized to thigh muscles, with decreased volume in 8/26 and increased IMF in 6/26. Acute pain groups showed no significant differences except for decreased volume in one thigh muscle in acute knee pain. With each additional chronic pain site, volume decreased (β=−.078;IQR:−0.100−0.051), and IMF increased (β=.085;IQR:0.066−0.101). Combined Z-scores classified chronic pain groups better than chance (accuracy: 48.6%;p<.001), but not acute pain groups (accuracy: 39.0%;p=.20).

**Conclusions:** Whole-body MRI combined with AI-driven muscle segmentation and normative modeling revealed site-specific patterns of muscle health in single-site chronic pain.

## 1. Introduction

Changes in skeletal muscle health are present in chronic pain conditions. Muscle atrophy and altered muscle composition are associated with poorer functional outcomes and greater pain persistence, suggesting that muscle health may play a causal role in sustaining chronic pain^1^. MRI is the gold standard for assessing muscle health, but its high costs often limit imaging to the primary site of pain. Consequently, few studies have investigated the effects of pain on muscle beyond the site of pain, and the mechanistic understanding of changes in muscle health—whether they are due to localized pain-site-specific factors, broader systemic factors, or their combination—remains unclear^2,3^. This knowledge gap limits our ability to distinguish between local and systemic drivers of pain, determine the clinical importance of changes in muscle health, and develop targeted interventions aimed at maintaining or improving muscle health.

The UK Biobank (UKB), the world’s largest prospective cohort study with both whole-body (neck-to-knees) MRI and pain measures in >50,000 people, provides an unprecedented opportunity to study muscular mechanisms of pain^4^. Historically, large-scale studies have been limited by the resource-intensive demands of manual muscle segmentation. However, recent advancements in computer vision, specifically artificial intelligence (AI)-powered image segmentation models, have overcome this barrier. This can enable the rapid, objective, and three-dimensional quantification of muscle size and composition at scale^5^. Despite automated segmentation and processing tools, an ongoing challenge is that muscle size and composition vary with age^6^, sex^7^, and body size^8^. If not accounted for, these factors can contribute to the misinterpretation of muscle health. Normative modeling is a framework that accounts for this variance, enabling patient-level inference and quantification of a person’s deviation from normal^9^. This approach has been extended to brain imaging, and normative brain markers have yielded larger effect sizes and better diagnostic performance in psychiatric and neurological conditions than raw, unadjusted imaging measures^10,11^. Here, we leverage the UKB, AI-powered muscle segmentation tools, and normative modeling to uncover whole-body muscular patterns of chronic pain at a large scale.

## 2. Materials and Methods

### 2.1 Dataset

This retrospective cross-sectional analysis used prospectively collected data from the UKB (Application 67450). The UK Biobank received approval from the National Information Governance Board for Health and Social Care and the National Health Service Northwest Multicentre Research Ethics Committee. All participants provided written informed consent prior to enrolment. The data used in this study are available to eligible researchers from academic, charity, government, and commercial organizations worldwide through the UK Biobank for health-related research in the public interest, subject to UK Biobank application and approval procedures. The analysis code supporting this work is publicly available online (https://github.com/NeuromuscularInsightLab/Muscle_Normative_Modeling). We included images from right-handed participants who attended the initial imaging assessment and underwent Dixon fat-water MRI. Imaging was performed in the supine position on a 1.5T Siemens Aera scanner using a multi-slab VIBE sequence (TE_1_/TE_2_=2.39/4.77ms, TR=6.69ms, flip angle=10°, in-plane resolution=2.232mm×2.232mm, slice thickness=4.5mm) covering the neck to knees (six overlapping slabs). The image slabs were stitched together into a single volume, the fat volume of which was input into the abdomen, pelvis, and thigh muscle segmentation models (v0.0) from the open-source MuscleMap Toolbox (https://musclemap.github.io/MuscleMap/^12^) to automatically segment 42 individual muscles (21 left[l]-right[r] pairs; Table S1). Using the segmentations, we assessed muscle size and composition by calculating muscle volume (ml) and intramuscular fat (IMF) for each muscle, respectively. IMF was calculated as the percentage of the total signal attributable to fat: (fat/(water+fat)×100). Two blinded raters (K.O. and K.A.W.) performed visual quality control of the images and segmentations. We excluded participants who withdrew consent; failed visual control; had missing data; were imaged at the Bristol imaging assessment center (<1% of dataset, UKB Data-Field 54); reported cancer, substance use disorder, or other major medical conditions (UKB Data-Fields 20001 and 20002); or had an age, height, or weight that fell outside ±3 standard deviations (SD) of the no pain group mean. In addition, participants in the no pain group were excluded if their muscle volume or IMF fell outside the no pain group mean ±3 SD to remove outliers prior to normative modeling.

We defined the no pain and site-specific pain groups using the pain questionnaires completed during the imaging assessment. Participants were asked whether in the last month they experienced “Headaches”, “Facial pain”, “Neck or shoulder pain”, “Back pain”, “Stomach or abdominal pain”, “Hip pain”, “Knee pain”, or “Pain all over the body”. Participants could choose multiple types of pain, except if “Pain all over the body” was selected. For each type of pain, participants were then asked if they had experienced pain for more than 3 months, which was used to define chronic (“Yes”) and acute (“No”) pain. Participants could also choose “None of the above” or “Prefer not to answer”. We defined the no pain group as a response of “None of the above”. We defined single-site chronic pain groups by identifying participants with only chronic neck/shoulder pain (CNSP), only chronic back pain (CBP), only chronic hip pain (CHP), or only chronic knee pain (CKP). Similarly, single-site acute pain groups were defined by identifying participants with only acute neck/shoulder pain (ANSP), only acute back pain (ABP), only acute hip pain (AHP), or only acute knee pain (AKP). Among participants with CBP, CHP, and/or CKP, we grouped participants by the number of chronic pain sites (CPS) across the back, hip, and knee: one site (1 CPS), two sites (2 CPS), or three sites (3 CPS). While the presence of acute pain at any site and of chronic pain at other body sites was allowed, only these three chronic pain sites were counted as CPS. Participants reporting “pain all over the body” were excluded to maintain a focus on localized chronic pain.

### 2.2. Normative Modeling

Using the no pain group as the reference cohort, we trained and tested hierarchical Bayesian regression (HBR) normative models for each muscle and measure (volume and IMF) using the PCNtoolkit (v1.1.2; https://github.com/amarquand/PCNtoolkit^13^). These normative models were then applied to the pain groups to compute muscle- and measure (volume and IMF)-specific Z-scores, quantifying how much an individual’s muscle volume and IMF deviate from the no pain group (Supplemental Material S1).

### 2.3 Statistical Analysis

All analyses were performed in Python (v3.11). Because muscle health measures in the chronic pain groups were skewed, with tails toward lower muscle volumes and higher IMF, median Z-scores were used as the measure of central tendency. To be robust to non-normal distributions and statistically more conservative, non-parametric tests were used for group-level analyses. Within each pain group and measure, we tested whether the median Z-score for each muscle deviated from zero (i.e., normal) using two-tailed sign-flip permutation tests, applying Bonferroni family-wise error (FWE) correction. For each measure and muscle, differences in median Z-scores between pair-wise pain groups were assessed using two-tailed permutation tests with Benjamini-Hochberg false discovery rate (FDR) correction. Linear trends in muscle Z-scores with increasing numbers of chronic pain sites were assessed using median (quantile) regression with group coded as an ordinal predictor and Benjamini–Hochberg FDR correction.

To test the predictive value of normative markers, we trained a logistic regression model using muscle volume and IMF Z-scores as features to classify between the single-site chronic pain groups. Performance was assessed using nested stratified cross-validation, with macro-F1 and balanced accuracy to account for class imbalance. Feature contributions were evaluated through iterative feature ablation analysis (see Supplemental Material S6). We similarly classified the single-site acute pain groups. Classification significance was evaluated using permutation testing.

All permutation-based analyses used 100,000 iterations; multiple-comparison correction was applied within each analysis across the 42 muscles, and significance was set at corrected p or q<.05.

## 3. Results

### 3.1 Participants

Of 34,786 right-handed UKB participants who underwent Dixon imaging at the initial imaging visit, 21,232 met the inclusion criteria after initial screening. We excluded 3,389 participants due to withdrawal from the UKB, missing measures, inadequate field-of-view, fat-water swapping artifacts, poor image quality, or poor segmentation quality, leaving 17,843 participants (Figure 1). The sample sizes for the pain and no pain groups were as follows: 9,704 no pain, 794 CNSP, 885 CBP, 438 CHP, 1,315 CKP, 413 ANSP, 667 ABP, 191 AHP, and 486 AKP. Additionally, 4,387 participants reported 1 CPS, 1,225 2 CPS, and 292 3 CPS. Participant characteristics are summarized in Table 1.

**Table 1.**
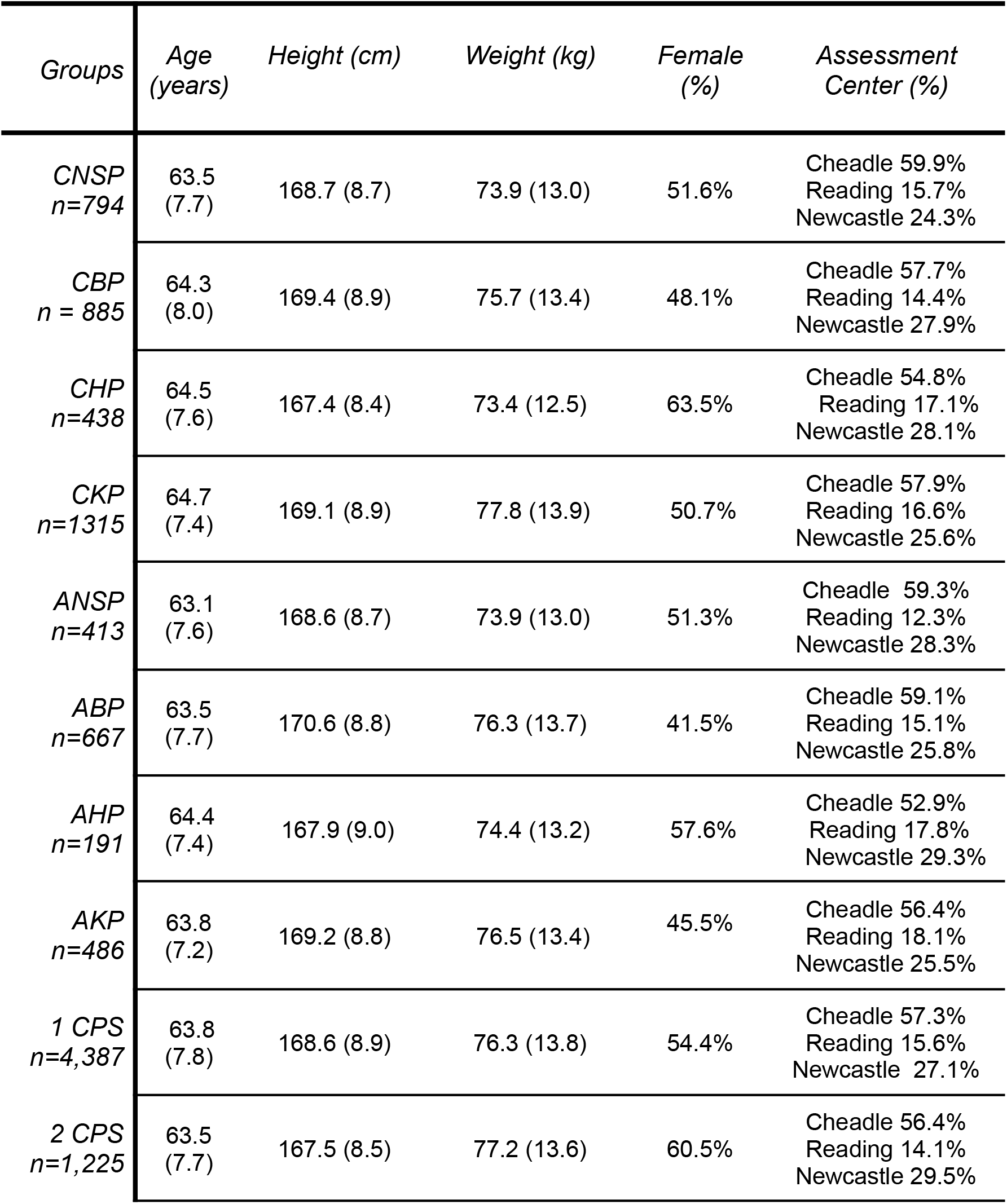
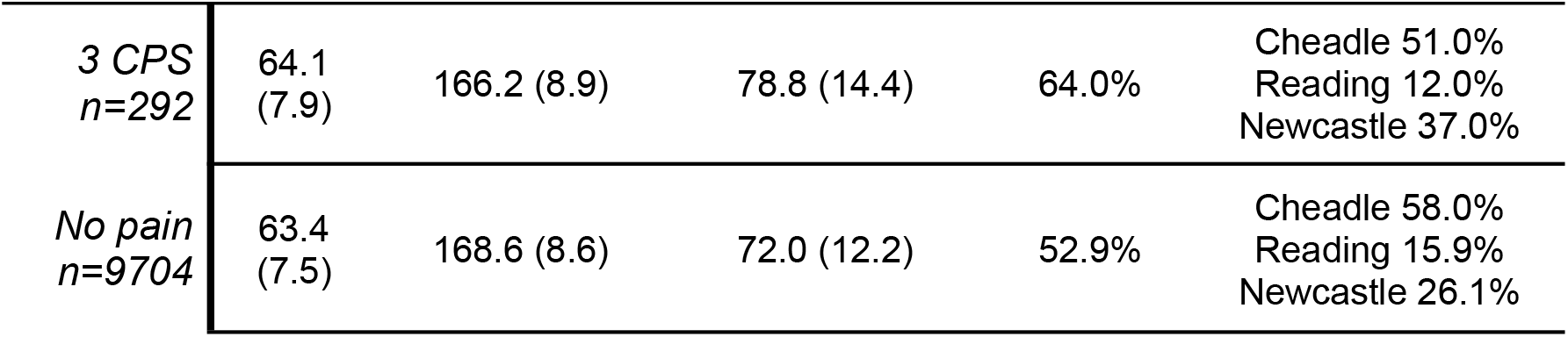
Demographic variables per group are presented as mean (SD) for age (years), height (cm), and weight (kg), and percentages for female sex and assessment centers.

**Figure 1.**
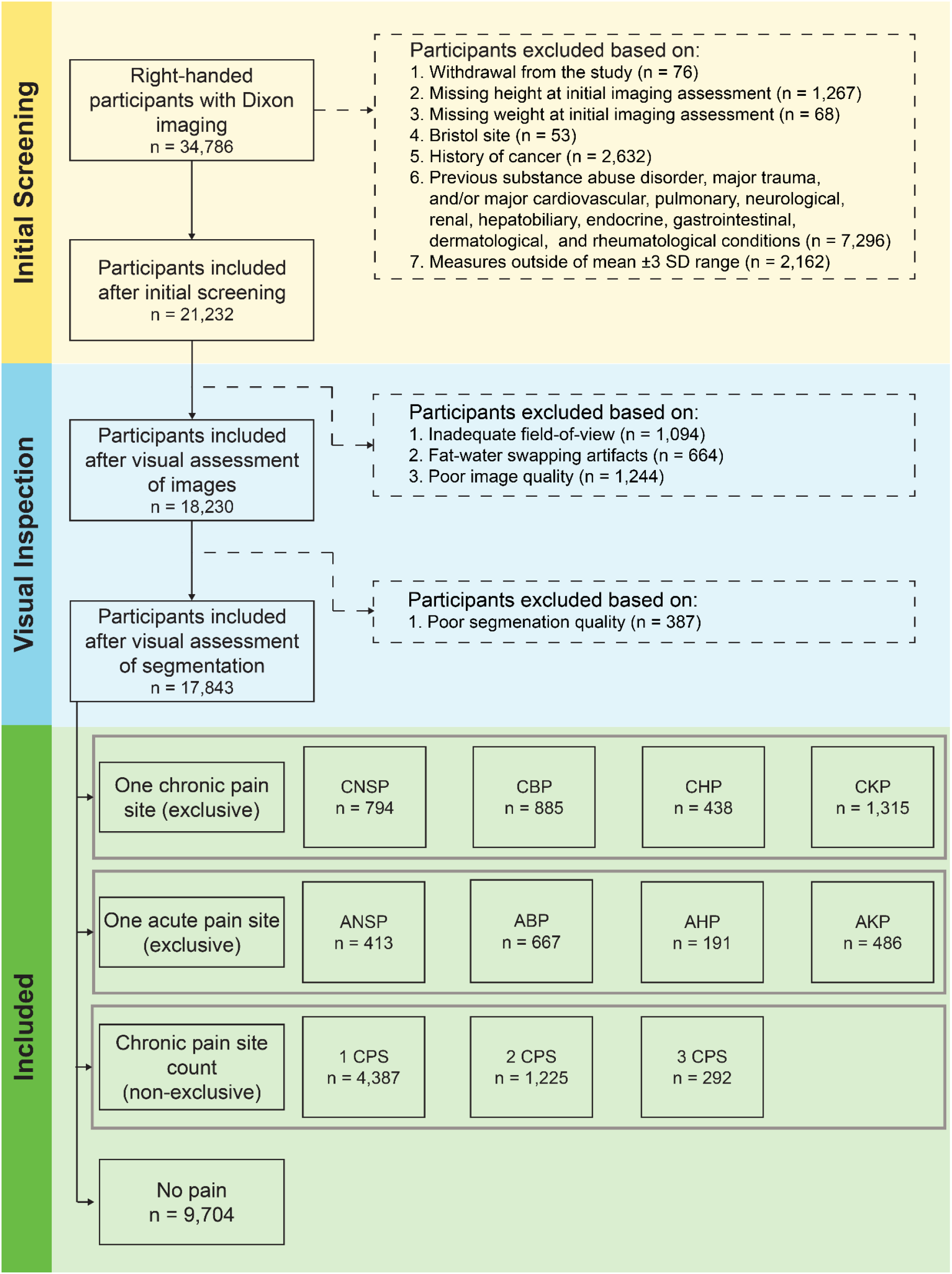
Flowchart for inclusion and exclusion of participants from the UK Biobank.

### 3.2 Normative Modeling Performance

Overall, muscle volume models outperformed IMF models. The best performing models for volume were observed for two bilateral thigh muscles (lVL and rVL) and one pelvic muscle (lGMed) and the worst performing models were one thigh muscle (lGra) and two bilateral pelvic muscles (rTFL and lTFL). The best-performing IMF models were observed in three abdominal muscles (lMF, rMF, lES), and the worst-performing were in two bilateral thigh muscles (lAdL and rAdB) and one abdominal muscle (lPM). Importantly, even the worst-performing models showed relatively stable performance across repeated train-test splits and remained within a reasonably good range across evaluation metrics, indicating that the normative models adequately captured the underlying distributions and produced consistent out-of-sample predictions (Supplemental Material S1).

### 3.3 Muscle Health Patterns in Single-Site Chronic Pain

Pain site-specific patterns in muscle health were observed in single-site chronic pain for both muscle volume and IMF (Figure 2). In CBP, the deviations in muscle health were more localized to the abdominal muscles. Decreased volume was seen in 75.0% (6/8) of abdominal muscles compared to 15.4% (4/26) of thigh muscles and no pelvic muscles. Increased IMF was seen in 75.0% (6/8) of abdomen muscles compared to 12.5% (1/8) of the pelvic muscles and no thigh muscles. In CHP, the deviations in muscle health were more broadly distributed. Decreased muscle volume was seen in 37.5% (3/8) of the abdominal and 53.8% (14/26) of the thigh muscles, but no pelvic muscles. Increased IMF was seen in 75.0% (6/8) of abdomen muscles, 62.5% (5/8) of the pelvic muscles, and 15.4% (4/26) of the thigh muscles. In CKP, the deviations were more localized to the thigh muscles, with bidirectional differences in muscle health. Decreased muscle volume was seen in 30.8% (8/26) of thigh muscles compared to 12.5% (1/8) of pelvic muscles and no abdominal muscles. Increased IMF was seen in 23.1% (6/26) of thigh muscles and no abdomen or pelvic muscles. Increased muscle volume was seen in 7.7% (2/26) of thigh muscles, and decreased IMF was seen in 11.5% (3/26) of thigh muscles and 12.5% of the pelvic muscles (1/8). In contrast, CNSP showed decreased volume in 23.1% (6/26) of thigh muscles, but no abdominal or pelvic muscles, and the magnitude of the decreases in volume was lower than that seen in CBP, CHP, and CKP. No IMF differences were observed in CNSP. See Table S2 for a full list of significant muscles. See Figures S2–S4 for between-group pairwise comparisons, which further highlight these pain site-specific patterns of muscle health in chronic pain.

**Figure 2.**
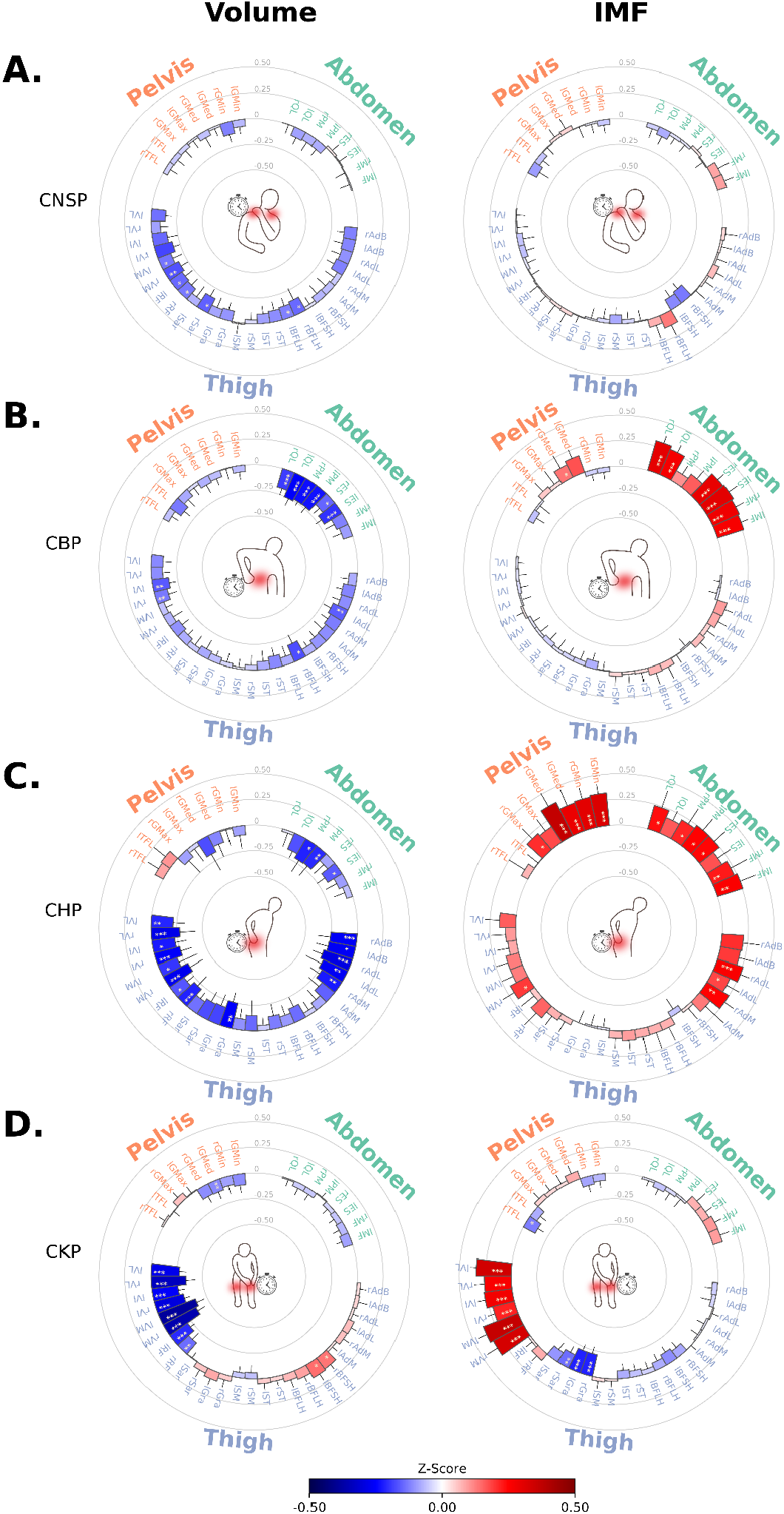
Patterns of muscle health in single-site chronic pain. Circle plots showing median normative Z-scores for muscle volume and intramuscular fat (IMF) across 42 abdomen, pelvis, and thigh muscles in people with single-site A) chronic neck and shoulder pain (CNSP), B) chronic back pain (CBP), C) chronic hip pain (CHP), and D) chronic knee pain (CKP). Error bars denote the 95% confidence interval. Outward bars (red) indicate higher than normal values; inward (blue) bars indicate lower than normal values. Asterisks denote muscles with group median Z-scores significantly differed from zero (* p<.05; ** p<.01; *** p<.001). **Abbreviations:** l=left; r=right. MF=multifidus; ES=erector spinae; PM=psoas major; QL=quadratus lumborum; GMin=gluteus minimus; GMed=gluteus medius; GMax=gluteus maximus; TFL=tensor fasciae latae; VL=vastus lateralis; VI=vastus intermedius; VM=vastus medialis; RF=rectus femoris; Sar=sartorius; Gra=gracilis; SM=semimembranosus; ST=semitendinosus; BFLH=biceps femoris long head; BFSH=biceps femoris short head; AdM=adductor magnus; AdL=adductor longus; AdB=adductor brevis.

### 3.4. Muscle Health Patterns in Single-Site Acute Pain

In ANSP, ABP, and AHP, no differences in muscle volume and IMF of the abdomen, pelvic, or thigh muscles were seen. In AKP, muscle volume was decreased in 3.8% (1/26) of thigh muscles (rVM; median:-0.19; 95% CI:-0.30–-0.03; p=.03) but not in any abdomen or pelvic muscles (Figure 3). Between-group pairwise comparisons show decreased volume and increased IMF in single-site chronic pain compared to single-site acute pain. None of the muscles demonstrated decreased volume or increased IMF in single-site acute pain compared to single-site chronic pain (Figure S5).

**Figure 3.**
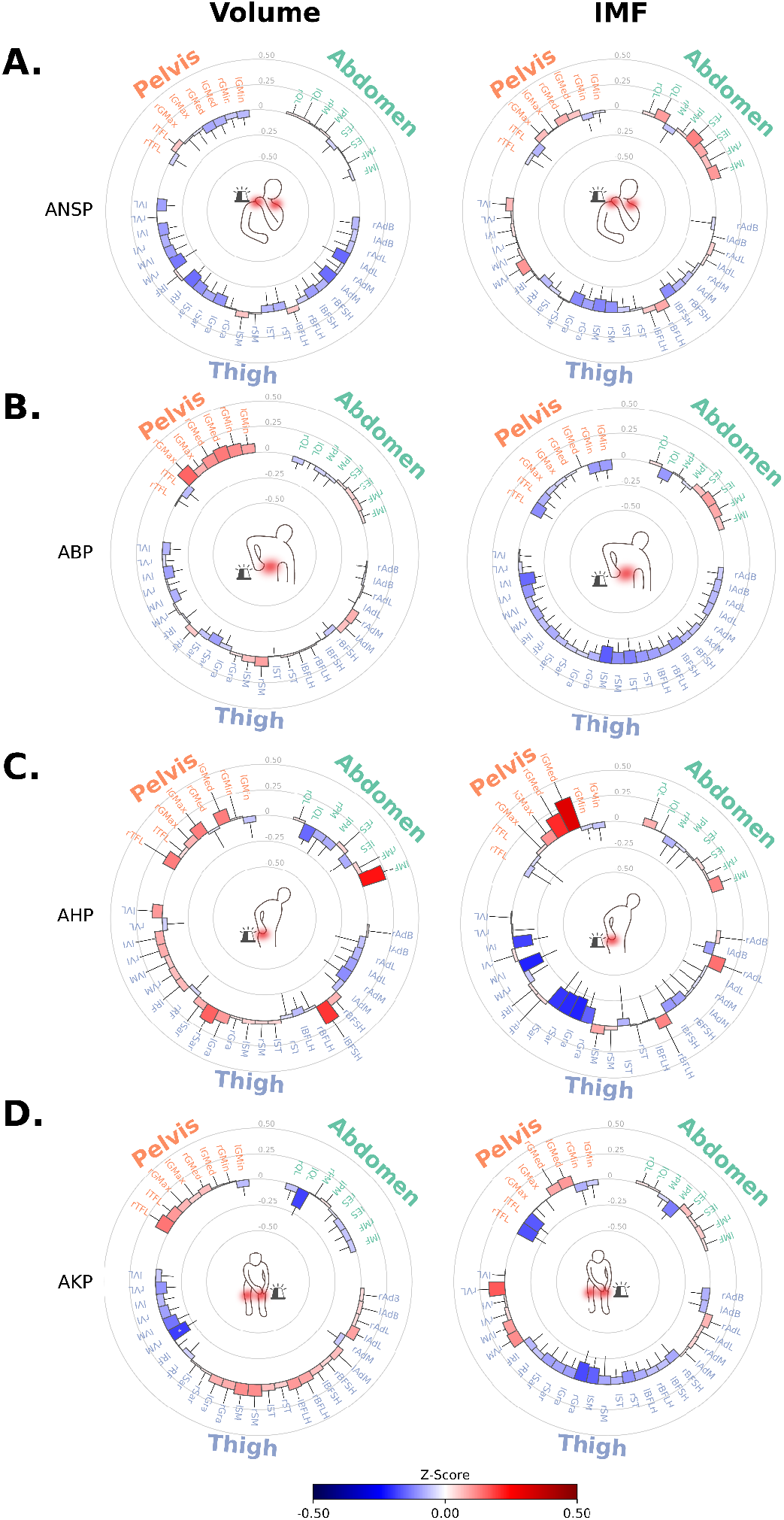
Patterns of muscle health in single-site acute pain. Circle plots showing median normative Z-scores for muscle volume and intramuscular fat (IMF) across 42 abdomen, pelvis, and thigh muscles in people with single-site, A) acute neck/shoulder pain (ANSP), B) acute back pain (ABP), C) acute hip pain (AHP), and D) acute knee pain (AKP). Error bars denote the 95% confidence interval. Outward bars (red) indicate higher than normal values; Inward (blue) bars indicate lower than normal values. Asterisks mark muscles with group median Z-scores significantly differed from zero (* p<.05; ** p<.01; *** p<.001). **Abbreviations:** l=left; r=right. MF=multifidus; ES=erector spinae; PM=psoas major; QL=quadratus lumborum; GMin=gluteus minimus; GMed=gluteus medius; GMax=gluteus maximus; TFL=tensor fasciae latae; VL=vastus lateralis; VI=vastus intermedius; VM=vastus medialis; RF=rectus femoris; Sar=sartorius; Gra=gracilis; SM=semimembranosus; ST=semitendinosus; BFLH=biceps femoris long head; BFSH=biceps femoris short head; AdM=adductor magnus; AdL=adductor longus; AdB=adductor brevis.

### 3.5. Patterns of Muscle Health with Increasing Number of CPS

Muscle volume decreased, and IMF increased with the increasing number of CPS (Figure 4). Median quantile regression demonstrated a consistent linear trend across groups for each muscle-outcome pair. All 42 muscles showed negative slopes for muscle volume and positive slopes for IMF, with 61.9% (26/42) and 66.7% (28/42) surviving FDR correction, respectively. The median slope across muscles was β=−0.078 (IQR=−.100–−.051) for muscle volume and β=0.085 (IQR=.066–.101) for IMF, for each additional CPS (Figure S6). See Table S3 for a full list of significant muscles.

**Figure 4.**
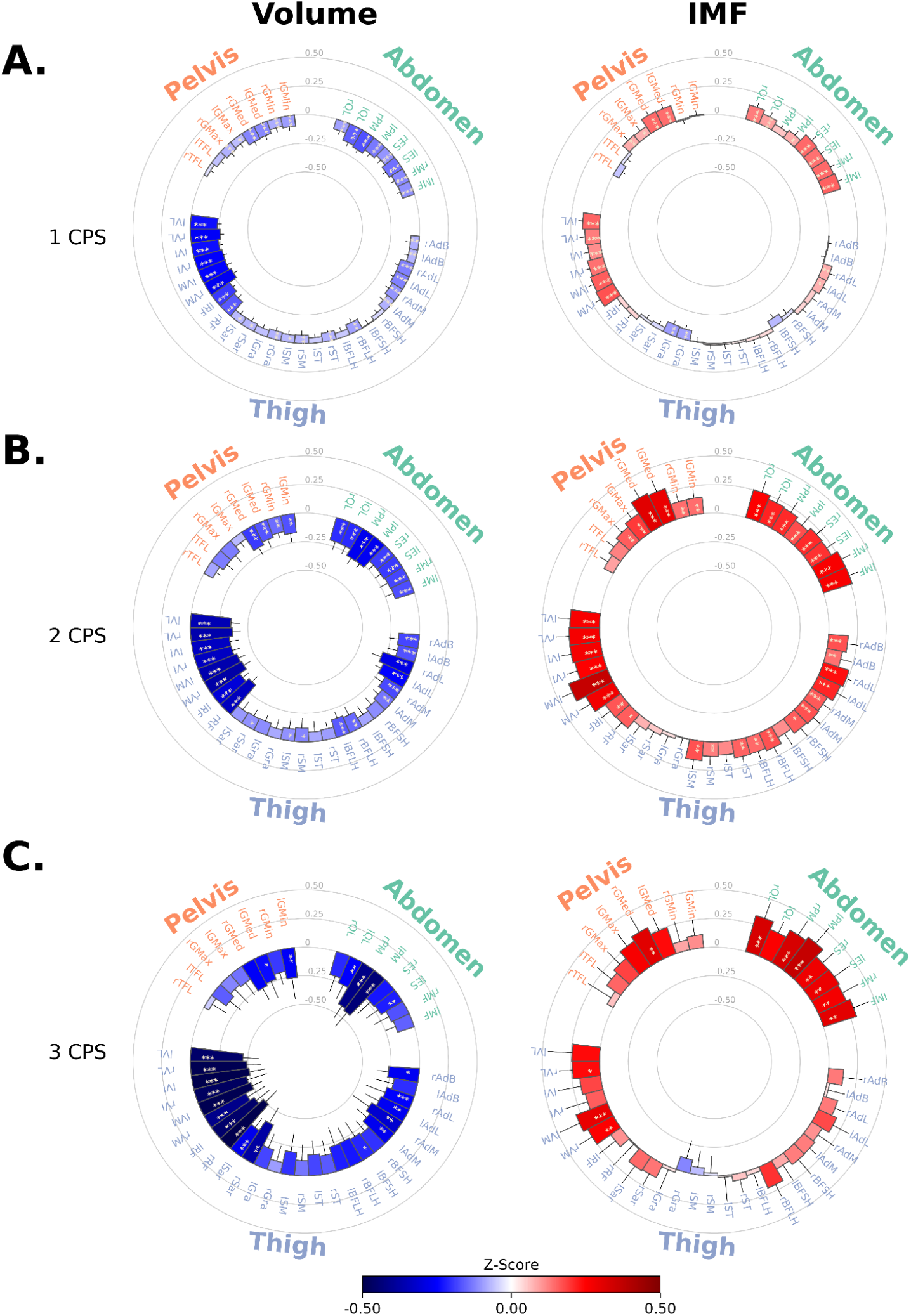
Patterns of muscle health with increasing number of chronic pain sites (CPS). Circle plots showing median normative Z-scores for muscle volume and intramuscular fat (IMF) across 42 abdomen, pelvis, and thigh muscles in people with chronic pain at A) 1 chronic pain site (CPS), B) 2 CPS, and C) 3 CPS. Error bars denote the 95% confidence interval. Outward bars (red) indicate higher than normal values; Inward (blue) bars indicate lower than normal values. Asterisks mark muscles with group median Z-scores significantly differed from zero (* p<.05; ** p<.01; *** p<.001). **Abbreviations:** l=left; r=right. MF=multifidus; ES=erector spinae; PM=psoas major; QL=quadratus lumborum; GMin=gluteus minimus; GMed=gluteus medius; GMax=gluteus maximus; TFL=tensor fasciae latae; VL=vastus lateralis; VI=vastus intermedius; VM=vastus medialis; RF=rectus femoris; Sar=sartorius; Gra=gracilis; SM=semimembranosus; ST=semitendinosus; BFLH=biceps femoris long head; BFSH=biceps femoris short head; AdM=adductor magnus; AdL=adductor longus; AdB=adductor brevis.

### 3.6. Predictive Value of Muscle Health Patterns

The combined muscle volume and IMF Z-scores were able to classify between the CBP, CHP, and CKP better than chance (macro-F1=47.4%, balanced accuracy=48.6%, p<.001, sensitivity=48.8%, specificity=74.9%, positive predictive value[PPV]=47.7%, negative predictive value[NPV]=74.4%). The features were not predictive of acute pain (macro-F1=37.2%, balanced accuracy=39.0%, p=.20, sensitivity=39.0%, specificity=69.8%, PPV=38.8%, NPV=69.5%). Feature importance analysis showed that the strongest predictive feature for classifying CBP, CHP, and CKP was a muscle corresponding to the site of pain (i.e., lPM, lGMin, and rVM, respectively; Figure 5).

**Figure 5.**
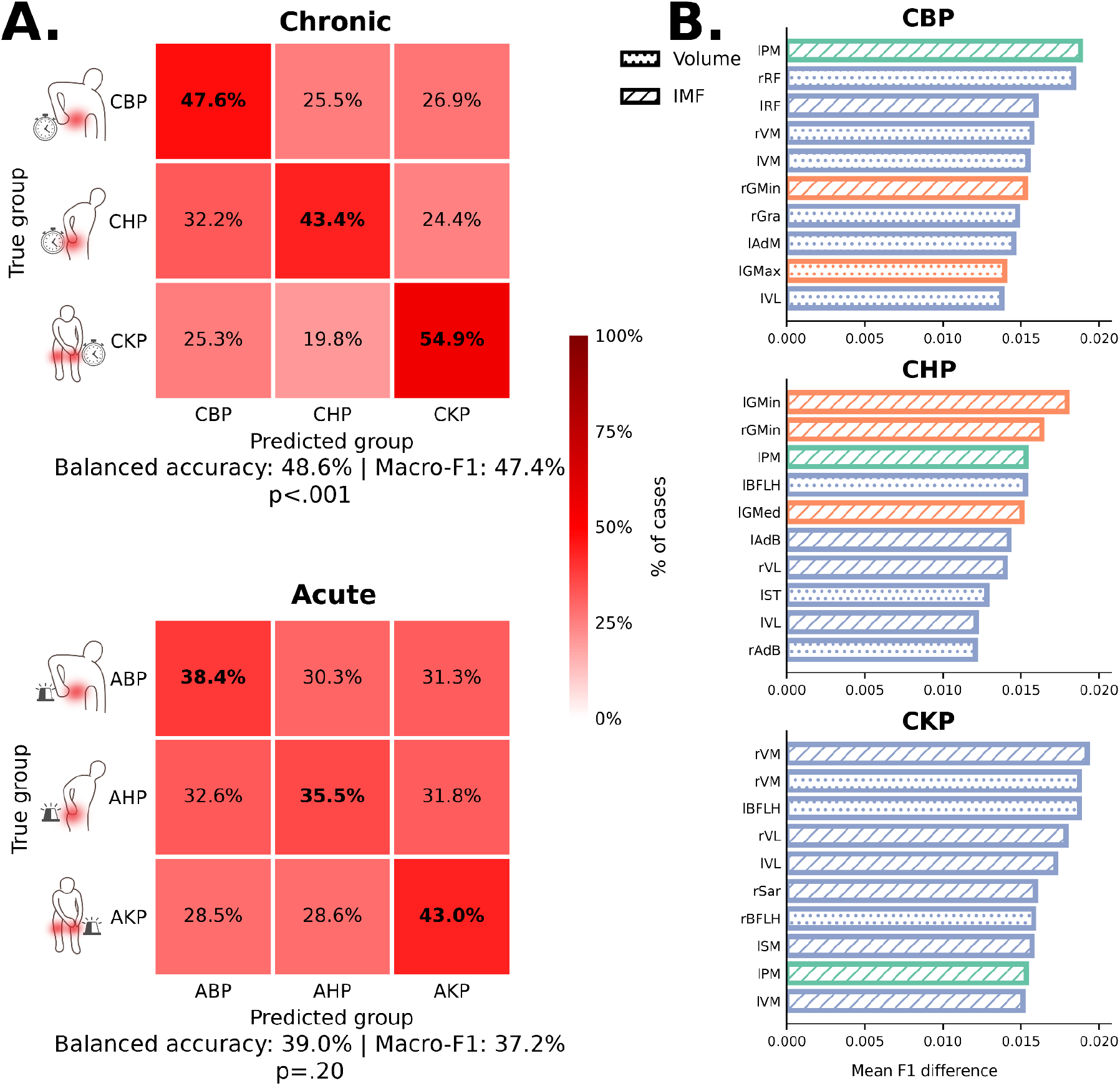
Predictive value of muscle health patterns in single-site chronic and acute pain. A) Confusion matrices show classification performance across pain groups for single-site chronic pain classification (chronic back pain: CBP, chronic hip pain: CHP; chronic knee pain: CKP), and single-site acute pain classification (acute back pain: ABP, acute hip pain: AHP; acute knee pain: AKP). In each confusion matrix, rows denote the true group labels and columns denote the predicted group labels. B) Bar plots (dotted for volume, striped for IMF) on the right display the 10 most important features contributing to chronic pain classification in each group. **Abbreviations:** l=left; r=right. MF=multifidus; ES=erector spinae; PM=psoas major; QL=quadratus lumborum; GMin=gluteus minimus; GMed=gluteus medius; GMax=gluteus maximus; TFL=tensor fasciae latae; VL=vastus lateralis; VI=vastus intermedius; VM=vastus medialis; RF=rectus femoris; Sar=sartorius; Gra=gracilis; SM=semimembranosus; ST=semitendinosus; BFLH=biceps femoris long head; BFSH=biceps femoris short head; AdM=adductor magnus; AdL=adductor longus; AdB=adductor brevis.

## 4. Discussion

Declines in skeletal muscle health are commonly observed in chronic pain conditions; however, the mechanistic understanding of these changes—whether they are due to localized pain-site-specific factors, more broad systemic factors, or their combination—remains unclear. We leveraged whole-body fat-water Dixon MRI from the UKB, AI-powered muscle segmentation tools, and normative modeling to study patterns of deviations in muscle health across multiple pain sites at a large scale. In people with single-site chronic pain, but not single-site acute pain, we provide strong evidence of site-specific patterns of muscle health, supporting more localized, pain-site-specific muscular mechanisms of chronic pain. In contrast, people with multi-site chronic pain showed broader patterns of deviations in muscle health, pointing towards the accumulative effects of multi-site pain on muscle health and/or other systemic mechanisms. Finally, we show that the normative markers of muscle health can classify between the site of pain in people with single-site chronic pain but not acute pain. Below, we discuss how these findings provide insight into potential mechanisms of chronic pain and how normative markers of muscle health, extracted from radiological workflows, could inform the clinical management of pain.

Site-specific patterns of muscle health in chronic pain may reflect a combination of impaired neural drive, altered biomechanics, and region-specific muscle adaptations. In CKP, quadriceps atrophy and increased quadriceps IMF alongside hamstring hypertrophy are consistent with the quadriceps-hamstring imbalance commonly reported in chronic knee conditions^14,15^. Two proposed contributors are arthrogenic muscle inhibition, which limits voluntary quadriceps activation^16^, and quadriceps avoidance gait, a compensatory strategy that reduces knee joint loading by minimizing quadriceps use^17,18^. In CBP, reduced volume and increased IMF in the lumbar paraspinal muscles are similarly consistent with local neural inhibition^19^ and pain-related avoidance behavior^20^. In contrast, CHP showed broader deviations from normal, which may reflect the hip’s central biomechanical role in linking trunk stability and lower-limb function^21–23^. Differences in muscle fiber type composition may also contribute, as Type I fiber-rich muscles, such as gluteus medius and minimus, showed larger IMF deviations than gluteus maximus, a Type II fiber-rich muscle, in CHP^24^. Finally, CNSP – included as a negative control – showed fewer and smaller deviations in muscle volume and no significant IMF deviations, supporting the specificity of these patterns to the site of pain. Notably, we did not observe deviations in muscle volume or IMF in ABP or AHP, and only one thigh muscle showed atrophy in AKP. This indicates that muscle changes may be absent or at least smaller in the early stages of pain (<3 months duration) and suggests that declines in muscle health evolve with pain duration, highlighting the potential value of early, site-specific rehabilitation and interventions to prevent pain-related declines in muscle health.

We observed an ordinal relationship between the number of chronic pain sites and deviations in muscle health, with muscle volume decreasing and IMF increasing. This dose-response pattern suggests that while single-site pain is associated with localized alterations, multi-site chronic pain is linked to broader and systemic muscle changes. This highlights a potential transition from regional adaptations to more generalized impairment with increasing pain burden, which may result from combined localized declines in muscle health and/or systemic factors (pain-related deconditioning, central sensitization, inflammation, and metabolic dysregulation), leading to broad changes in muscle health.

The normative modeling framework developed here is analogous to DEXA-based bone densitometry, where individual deviations from population norms guide clinical decision-making for osteoporosis. Similarly, muscle health Z-scores could serve as quantitative biomarkers in radiology. Because muscle is already captured in most clinical MRI and CT examinations, AI-powered tools such as the MuscleMap Toolbox could extract these normative markers without additional scan time or cost. In surgical settings, such profiles could inform prehabilitation, set outcome expectations, and support candidacy decisions, while longitudinal tracking could quantify treatment response. Clinical translation will require prospective validation, cross-scanner generalization, integration into PACS-based workflows, and establishment of clinically meaningful Z-score thresholds. The normative models and calibration scripts will be released in the next MuscleMap Toolbox update to support these efforts.

This study has several limitations. While the normative markers could classify single-site chronic pain better than chance, their accuracy was low and of limited predictive value, indicating that muscle health patterns are complex and variable within and between people with CBP, CHP, and CKP. Pain-related variables available at the imaging assessment were also limited, preventing further evaluation of associations between normative muscle markers and pain intensity, duration, and disability. We also assessed only a subset of muscles due to image resolution, field of view, and current constraints in automated segmentation. Additionally, the cross-sectional design precludes causal inference. Finally, the normative models were developed in UKB using a limited set of covariates and standardized Siemens acquisitions, requiring external validation and recalibration.

In this cross-sectional, large-scale UKB study, whole-body MRI combined with AI-driven muscle segmentation and normative modeling revealed site-specific patterns of muscle atrophy and increased IMF in single-site chronic pain that were absent in acute pain, with deviations broadening as the number of chronic pain sites increased. These normative markers, conceptually analogous to bone densitometry Z-scores, could provide a framework for quantifying individual-level muscle health from imaging already acquired in clinical practice.

## Data Availability

The data used in this study are available to eligible researchers from academic, charity, government, and commercial organizations worldwide through the UK Biobank for health-related research in the public interest, subject to UK Biobank application and approval procedures. The analysis code supporting this work is publicly available online (https://github.com/NeuromuscularInsightLab/Muscle_Normative_Modeling).

## 5. Acknowledgments

Kenneth A. Weber II received funding from the National Institute of Neurological Disorders and Stroke (grants R01NS133305 and R01NS128478). Yiyu Wang and Joel Fundaun are supported by the National Institute on Drug Abuse T32DA035165. Sandrine Bedard is supported by the Canada Graduate Scholarships-Doctoral program from the NSERC of Canada. Dario Pfyffer was supported by the Swiss National Science Foundation Postdoc Mobility Fellowship grant (P500PM_214211). Julien Cohen-Adad is funded by the Canada Research Chair in Quantitative Magnetic Resonance Imaging [CRC-2020-00179], the Canadian Institute of Health Research [PJT-190258, PJT-203803], the Canada Foundation for Innovation [32454, 34824], the Fonds de Recherche du Québec-Santé [322736, 324636], the Natural Sciences and Engineering Research Council of Canada [RGPIN-2019-07244, RGPIN-2025-05074], the Canada First Research Excellence Fund (IVADO and TransMedTech), Mila - Tech Transfer Funding Program. Andrew C. Smith was supported by the Eunice Kennedy Shriver National Institute of Child Health and Human Development of the National Institutes of Health – K01HD106928 and the Boettcher Foundation’s Webb-Waring Biomedical Research Program. The content is solely the responsibility of the authors and does not necessarily represent the official views of the National Institutes of Health. Brian Kim is supported by the University of Sydney Postgraduate Award and the Australian Government Department of Education Research Training Program. James M Elliott, Marnee McKay, Brian Kim, Eddo Wesselink, and Kenneth A. Weber II received philanthropic funding from Helena Charitable Foundation, Switzerland.

## 7. Ethics Statement

The UK Biobank received approval from the National Information Governance Board for Health and Social Care and the National Health Service Northwest Multicentre Research Ethics Committee. All participants provided written informed consent prior to enrolment.

