## Supplemental Material for "Patterns of Muscle Health in Single- and Multi-Site Chronic Pain: A UK Biobank Normative Modeling Study"

**Table S1.** Muscle abbreviations and body regions for the 21 bilateral muscle pairs are automatically segmented by the MuscleMap Toolbox.

| Muscle | Abbreviation | Body Region |
|---|---|---|
| *Multifidus* | MF | Abdomen |
| *Erector Spinae* | ES | Abdomen |
| *Psoas Major* | PM | Abdomen |
| *Quadratus Lumborum* | QL | Abdomen |
| *Gluteus Minimus* | GMin | Pelvis |
| *Gluteus Medius* | GMed | Pelvis |
| *Gluteus Maximus* | GMax | Pelvis |
| *Tensor Fasciae Latae* | TFL | Pelvis |
| *Vastus Medialis* | VM | Thigh |
| *Vastus Intermedius* | VI | Thigh |
| *Vastus Lateralis* | VL | Thigh |
| *Rectus Femoris* | RF | Thigh |
| *Sartorius* | Sar | Thigh |
| *Gracilis* | Gra | Thigh |
| *Semimembranosus* | SM | Thigh |
| *Semitendinosus* | ST | Thigh |
| *Biceps Femoris (Long Head)* | BFLH | Thigh |
| *Biceps Femoris (Short Head)* | BFSH | Thigh |
| *Adductor Magnus* | AdM | Thigh |
| *Adductor Longus* | AdL | Thigh |
| *Adductor Brevis* | AdB | Thigh |

### S1. Normative Modeling

Muscle volume and IMF were modeled with cubic B-spline expansions of age, height, and weight (five knots), additive random intercepts for sex and assessment center, and a covariate-dependent linear variance term to account for heteroscedasticity. The reference cohort was split 80%/20% (stratified by sex and assessment center), and out-of-sample performance was evaluated across 10 train-test splits.

We assessed the fit of normative models using quantitative out-of-sample diagnostics. Within a reference run, which was used to present the results in the main manuscript, we first identified the ‘best’ and ‘worst’ performing muscles for each measure (volume and IMF). We utilized three standard and complementary metrics to assess different aspects of the modeling: 1) Mean standardized log-loss (MSLL) as it is sensitive to both the predicted mean and the estimated predictive variance, 2) standardized mean squared error (SMSE) to quantify the accuracy of the predicted central tendency, and 3) Shapiro-Wilk test statistic (Shapiro-W) to assess whether the Z-scores follow a Gaussian distribution as a proxy metric for model calibration [1].

We then ranked each muscle within each measure using a composite score defined as the mean of three within-measure rank orders: MSLL rank (ascending; lower is better), SMSE rank (ascending; lower is better), and Shapiro-W rank (descending; higher is better). We present the three best- and three worst-performing muscles for each measure and demonstrate that their performance was relatively consistent across 10 independent train-test splits rather than being driven by a single model instance (Figure S1).

Across the selected muscles and repeated train-test splits, pooled performance summaries suggest better overall performance for muscle volume models than for IMF models, with muscle volume models showing more favorable MSLL, lower SMSE, and higher Shapiro-W values. For muscle volume, pooled MSLL was -5.437 ± 0.402 in the best-performing model and -3.050 ± 0.260 in the worst-performing model; corresponding SMSE values were 0.217 ± 0.018 and 0.479 ± 0.081, and Shapiro-W values were 0.996 ± 0.002 and 0.984 ± 0.003, respectively. For IMF, pooled MSLL was -2.427 ± 0.094 in the best-performing model and -0.742 ± 0.067 in the worst-performing model; corresponding SMSE values were 0.607 ± 0.016 and 0.796 ± 0.045, and Shapiro-W values were 0.992 ± 0.005 and 0.961 ± 0.007, respectively (Figure S1).

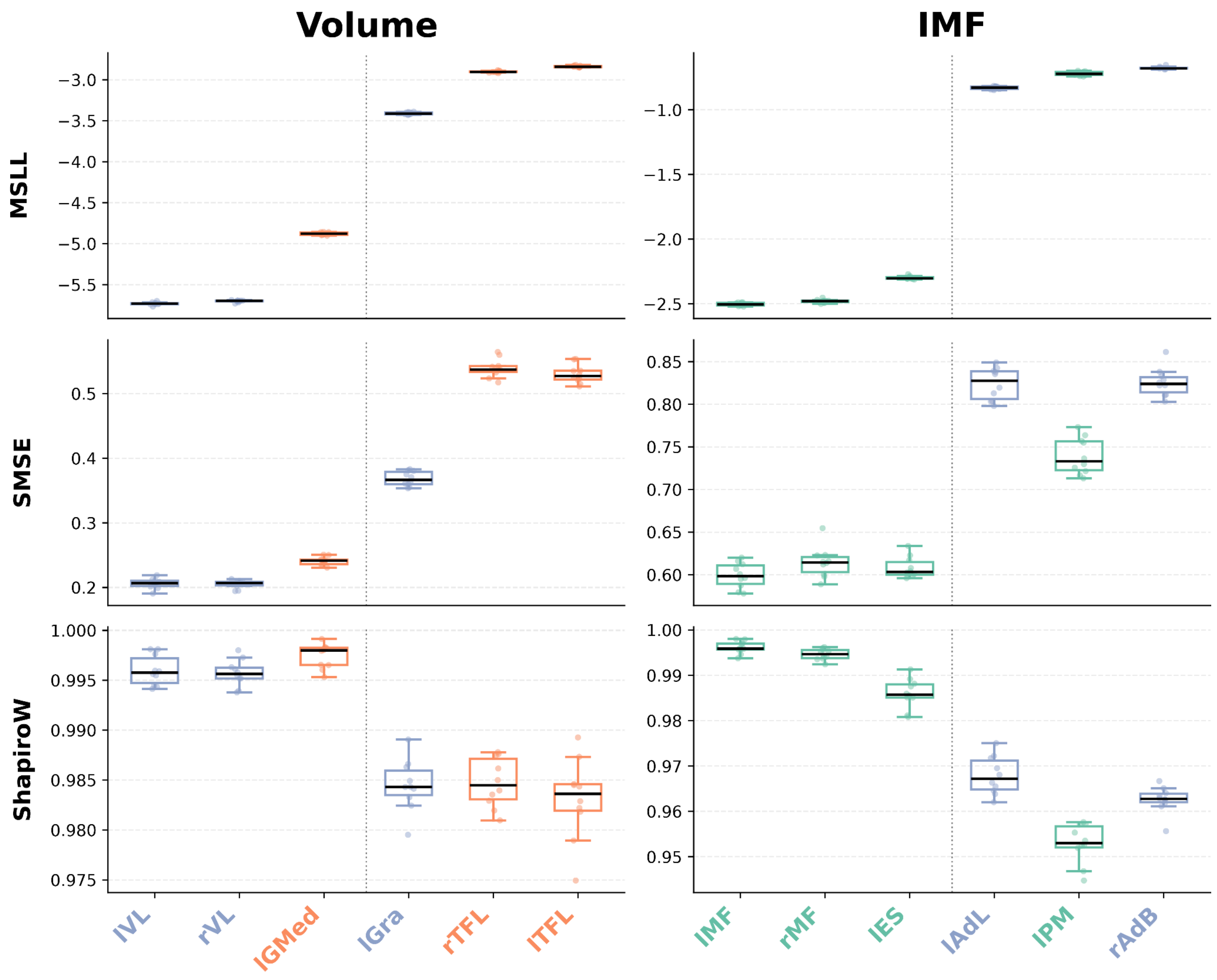


**Figure S1. Assessment of normative model performance and stability.** Boxplots summarize the distribution of held-out test performance metrics across 10 independent HBR model runs for the three best (left of the dotted line) and three worst (right of the dotted line) muscles selected separately for IMF (left column) and volume (right column) using a composite rank (average of muscle-wise ranks for MSLL and SMSE and Shapiro-W). Rows show MSLL (top), SMSE (middle), and Shapiro-W(bottom). Points denote individual runs; box centers and whiskers indicate median and interquartile range with 1.5×IQR whiskers. Muscle label colors indicate anatomical region (abdomen–blue, pelvis–orange, and thigh–green). **Abbreviations:** Mean standardized log-loss=MSLL; standardized mean squared error=SMSE; Shapiro-Wilk test statistic=Shapiro-W.

#### S2. Normative Modeling Results for Single-Site Chronic Pain

**Table S2. Deviations of muscle volume and IMF from normal in single-site chronic musculoskeletal pain.** The table shows all significant results derived from the normative modeling analysis for single-site chronic musculoskeletal pain groups. For each group and outcome (volume, IMF), the muscle abbreviation, median deviation, 95% confidence interval, and Bonferroni-corrected p-value are listed. Only results surviving Bonferroni-corrected significance threshold ($p < .05$) are shown. **Abbreviations:** l=left; r=right. MF=multifidus; ES=erector spinae; PM=psoas major; QL=quadratus lumborum; GMin=gluteus minimus; GMed=gluteus medius; GMax=gluteus maximus; TFL=tensor fasciae latae; VL=vastus lateralis; VI=vastus intermedius; VM=vastus medialis; RF=rectus femoris; Sar=sartorius; Gra=gracilis; SM=semimembranosus; ST=semitendinosus; BFLH=biceps femoris long head; BFSH=biceps femoris short head; AdM=adductor magnus; AdL=adductor longus; AdB=adductor brevis.

| Single-Site Chronic Musculoskeletal Pain | | | | | |
|---|---|---|---|---|---|
| **Group** | **Outcome** | **Muscle** | **Median Deviation** | **95% CI** | **p (corr.)** |
| CNSP | Volume | rVM | -0.17 | -0.25 – -0.07 | 0.003 |
| CNSP | Volume | lVM | -0.13 | -0.23 – -0.05 | 0.015 |
| CNSP | Volume | lRF | -0.14 | -0.24 – -0.06 | 0.033 |
| CNSP | Volume | rRF | -0.13 | -0.22 – -0.06 | 0.039 |
| CNSP | Volume | lBFLH | -0.12 | -0.21 – -0.06 | 0.039 |
| CNSP | Volume | rSar | -0.16 | -0.23 – -0.09 | 0.041 |
| CBP | Volume | lES | -0.20 | -0.27 – -0.10 | <.001 |
| CBP | Volume | lPM | -0.23 | -0.30 – -0.15 | <.001 |
| CBP | Volume | rPM | -0.23 | -0.28 – -0.17 | <.001 |
| CBP | Volume | lQL | -0.25 | -0.33 – -0.20 | <.001 |
| CBP | Volume | rQL | -0.19 | -0.25 – -0.11 | <.001 |
| CBP | Volume | lVI | -0.16 | -0.23 – -0.08 | 0.002 |
| CBP | Volume | lAdL | -0.16 | -0.24 – -0.11 | 0.003 |
| CBP | Volume | rVI | -0.13 | -0.21 – -0.06 | 0.008 |
| CBP | Volume | rBFLH | -0.17 | -0.26 – -0.06 | 0.014 |
| CBP | Volume | rES | -0.15 | -0.22 – -0.04 | 0.018 |
| CBP | IMF | lMF | 0.26 | 0.18 – 0.34 | <.001 |
| CBP | IMF | rMF | 0.28 | 0.19 – 0.37 | <.001 |
| CBP | IMF | lES | 0.30 | 0.24 – 0.40 | <.001 |

| | | | | | |
|---|---|---|---|---|---|
| CBP | IMF | rES | 0.29 | 0.20 – 0.37 | <.001 |
| CBP | IMF | lQL | 0.27 | 0.21 – 0.35 | <.001 |
| CBP | IMF | rQL | 0.30 | 0.20 – 0.38 | <.001 |
| CBP | IMF | lGMed | 0.17 | 0.04 – 0.26 | 0.046 |
| CHP | Volume | rVL | -0.28 | -0.43 – -0.15 | <.001 |
| CHP | Volume | rVI | -0.26 | -0.36 – -0.11 | <.001 |
| CHP | Volume | rVM | -0.27 | -0.42 – -0.16 | <.001 |
| CHP | Volume | rRF | -0.22 | -0.34 – -0.11 | <.001 |
| CHP | Volume | rAdL | -0.29 | -0.40 – -0.18 | <.001 |
| CHP | Volume | rAdB | -0.26 | -0.38 – -0.12 | <.001 |
| CHP | Volume | lPM | -0.21 | -0.33 – -0.08 | 0.003 |
| CHP | Volume | rAdM | -0.23 | -0.34 – -0.14 | 0.003 |
| CHP | Volume | lAdL | -0.26 | -0.43 – -0.19 | 0.003 |
| CHP | Volume | rGra | -0.25 | -0.34 – -0.13 | 0.004 |
| CHP | Volume | lVL | -0.21 | -0.33 – -0.13 | 0.008 |
| CHP | Volume | rPM | -0.22 | -0.34 – -0.07 | 0.01 |
| CHP | Volume | lRF | -0.17 | -0.25 – -0.06 | 0.024 |
| CHP | Volume | lVM | -0.2 | -0.39 – -0.03 | 0.031 |
| CHP | Volume | lVI | -0.22 | -0.33 – -0.07 | 0.034 |
| CHP | Volume | lES | -0.16 | -0.26 – -0.07 | 0.034 |
| CHP | Volume | lAdB | -0.26 | -0.38 – -0.10 | 0.045 |
| CHP | IMF | lGMin | 0.32 | 0.18 – 0.41 | <.001 |
| CHP | IMF | rGMin | 0.30 | 0.16 – 0.41 | <.001 |
| CHP | IMF | lGMed | 0.29 | 0.11 – 0.46 | <.001 |
| CHP | IMF | rGMed | 0.36 | 0.26 – 0.52 | <.001 |
| CHP | IMF | rAdL | 0.26 | 0.13 – 0.35 | <.001 |
| CHP | IMF | lMF | 0.26 | 0.14 – 0.41 | 0.003 |
| CHP | IMF | rMF | 0.23 | 0.09 – 0.40 | 0.008 |
| CHP | IMF | rAdM | 0.25 | 0.10 – 0.36 | 0.009 |

| | | | | | |
|---|---|---|---|---|---|
| CHP | IMF | rES | 0.24 | 0.14 – 0.38 | 0.013 |
| CHP | IMF | rPM | 0.21 | 0.10 – 0.39 | 0.013 |
| CHP | IMF | lPM | 0.25 | 0.09 – 0.40 | 0.015 |
| CHP | IMF | rQL | 0.24 | 0.12 – 0.41 | 0.02 |
| CHP | IMF | rGMax | 0.21 | 0.08 – 0.34 | 0.028 |
| CHP | IMF | lAdL | 0.18 | 0.08 – 0.31 | 0.042 |
| CHP | IMF | rVM | 0.19 | 0.09 – 0.32 | 0.043 |
| CKP | Volume | lVL | -0.27 | -0.34 – -0.19 | <.001 |
| CKP | Volume | rVL | -0.34 | -0.43 – -0.27 | <.001 |
| CKP | Volume | lVI | -0.25 | -0.31 – -0.18 | <.001 |
| CKP | Volume | rVI | -0.29 | -0.35 – -0.22 | <.001 |
| CKP | Volume | lVM | -0.39 | -0.47 – -0.31 | <.001 |
| CKP | Volume | rVM | -0.36 | -0.45 – -0.29 | <.001 |
| CKP | Volume | lRF | -0.21 | -0.27 – -0.14 | <.001 |
| CKP | Volume | rRF | -0.14 | -0.23 – -0.07 | 0.003 |
| CKP | Volume | lGMed | -0.11 | -0.18 – -0.03 | 0.007 |
| CKP | Volume | lBFSH | 0.14 | 0.06 – 0.22 | 0.01 |
| CKP | Volume | rBFSH | 0.13 | 0.06 – 0.19 | 0.013 |
| CKP | IMF | lVL | 0.34 | 0.22 – 0.40 | <.001 |
| CKP | IMF | rVL | 0.26 | 0.18 – 0.36 | <.001 |
| CKP | IMF | lVI | 0.27 | 0.17 – 0.35 | <.001 |
| CKP | IMF | rVI | 0.22 | 0.12 – 0.29 | <.001 |
| CKP | IMF | lVM | 0.36 | 0.26 – 0.45 | <.001 |
| CKP | IMF | rVM | 0.33 | 0.23 – 0.45 | <.001 |
| CKP | IMF | lGra | -0.21 | -0.27 – -0.13 | <.001 |
| CKP | IMF | rGra | -0.22 | -0.28 – -0.14 | <.001 |
| CKP | IMF | rSar | -0.14 | -0.21 – -0.07 | .008 |
| CKP | IMF | rTFL | -0.13 | -0.19 – -0.06 | .02 |

### S3. Pairwise Group Differences Between Single-Site Chronic Pain Groups

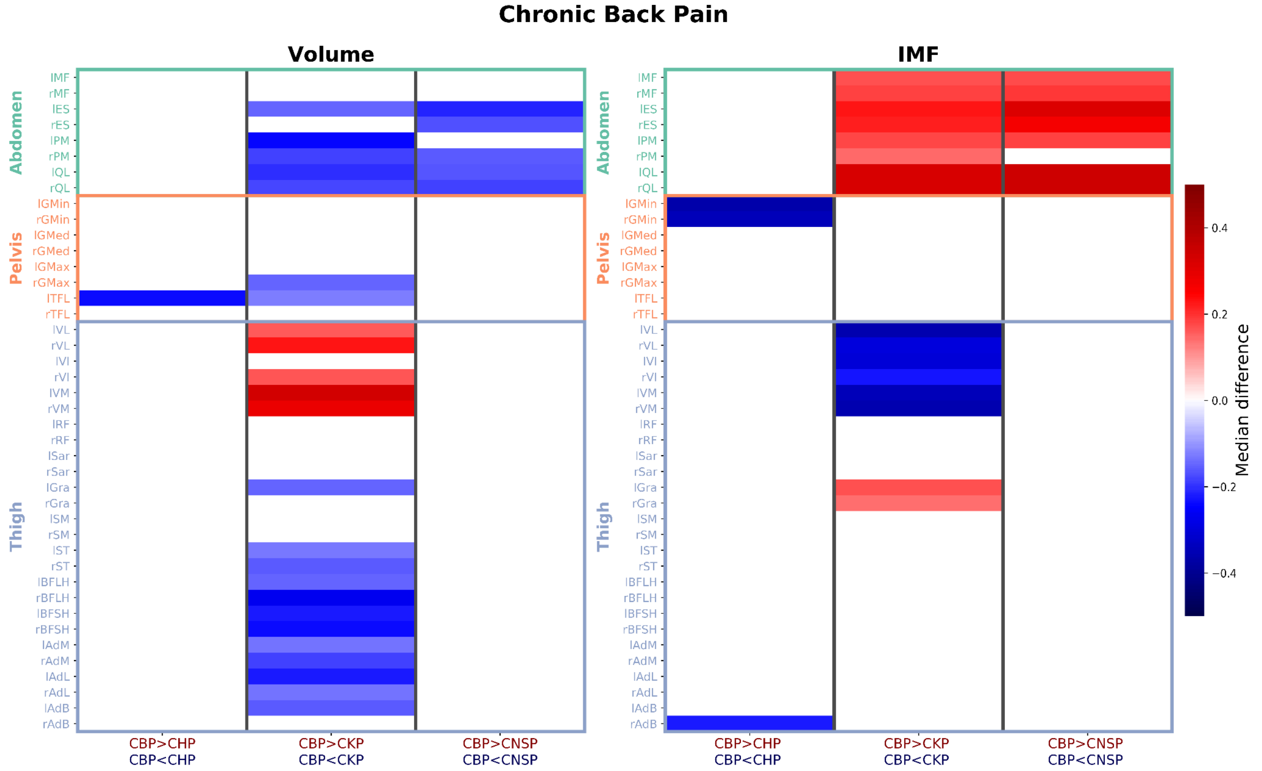


**Figure S2. Pairwise group differences in muscle Z-scores.** The heatmap shows median differences in normative Z-scores comparing participants with CBP to other chronic pain groups. Rows correspond to 42 bilateral muscle ROIs, grouped by region. Columns show the pairwise comparisons against the CBP group (CBP vs CHP, CBP vs CKP, CBP vs CNSP). Cell color denotes the median Z-score difference for each comparison, restricted to results surviving Benjamini–Hochberg FDR correction applied within each pairwise comparison across 42 muscles (q <.05); non-significant cells are shown in white. Red indicates CBP > comparison group and blue indicates CBP<comparison group. **Abbreviations:** CBP=Chronic Back Pain; CHP=Chronic Hip Pain; CKP=Chronic Knee Pain; CNSP =Chronic Neck/Shoulder Pain; l=left; r=right; MF=multifidus; ES=erector spinae; PM=psoas major; QL=quadratus lumborum; GMin=gluteus minimus; GMed=gluteus medius; GMax=gluteus maximus; TFL=tensor fasciae latae; VL=vastus lateralis; VI=vastus intermedius; VM=vastus medialis; RF=rectus femoris; Sar=sartorius; Gra=gracilis; SM=semimembranosus; ST=semitendinosus; BFLH=biceps femoris long head; BFSH=biceps femoris short head; AdM=adductor magnus; AdL=adductor longus; AdB=adductor brevis.

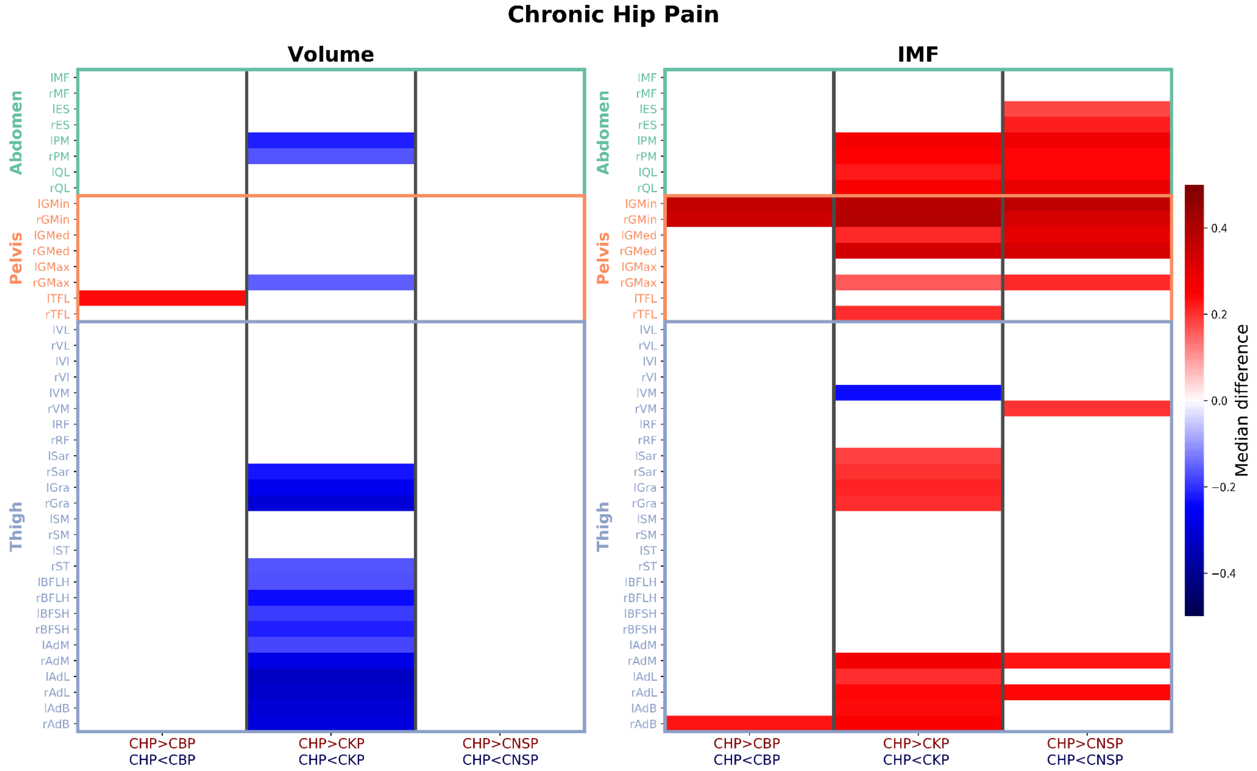


**Figure S3. Pairwise group differences in muscle Z-scores.** The heatmap shows median differences in normative Z-scores comparing participants with CHP to other chronic pain groups. Rows correspond to 42 bilateral muscle ROIs, grouped by region. Columns show the pairwise comparisons against the CHP group (CHP vs CBP, CHP vs CKP, CHP vs CNSP). Cell color denotes the median Z-score difference for each comparison, restricted to results surviving Benjamini–Hochberg FDR correction applied within each pairwise comparison across 42 muscles (q <.05); non-significant cells are shown in white. Red indicates CHP > comparison group and blue indicates CHP <comparison group. **Abbreviations:** CHP=Chronic Hip Pain; CBP=Chronic Back Pain; CKP=Chronic Knee Pain; CNSP =Chronic Neck/Shoulder Pain; l=left; r=right; MF=multifidus; ES=erector spinae; PM=psoas major; QL=quadratus lumborum; GMin=gluteus minimus; GMed=gluteus medius; GMax=gluteus maximus; TFL=tensor fasciae latae; VL=vastus lateralis; VI=vastus intermedius; VM=vastus medialis; RF=rectus femoris; Sar=sartorius; Gra=gracilis; SM=semimembranosus; ST=semitendinosus; BFLH=biceps femoris long head; BFSH=biceps femoris short head; AdM=adductor magnus; AdL=adductor longus; AdB=adductor brevis.

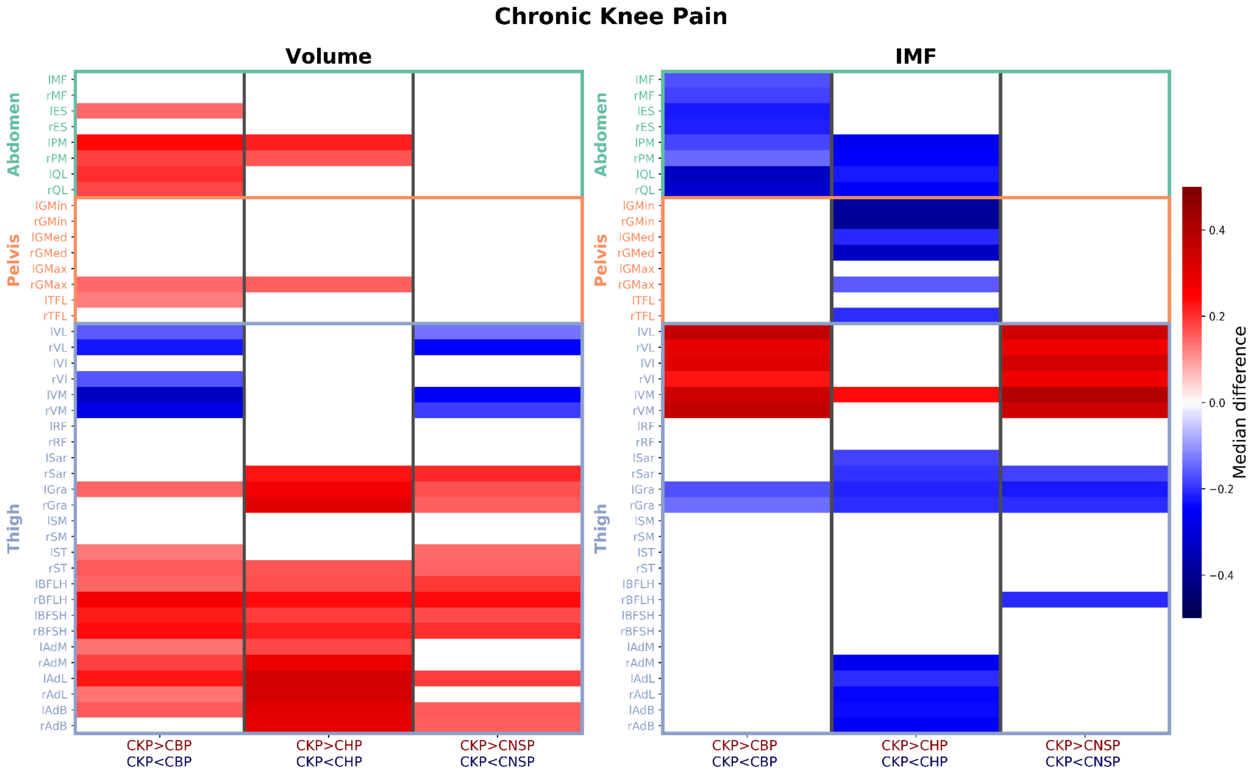


**Figure S4. Pairwise group differences in muscle Z-scores.** The heatmap shows median differences in normative Z-scores comparing participants with CKP to other chronic pain groups. Rows correspond to 42 bilateral muscle ROIs, grouped by region. Columns show the pairwise comparisons against the CKP group (CKP vs CBP, CKP vs CHP, CKP vs CNSP). Cell color denotes the median Z-score difference for each comparison, restricted to results surviving Benjamini–Hochberg FDR correction applied within each pairwise comparison across 42 muscles (q <.05); non-significant cells are shown in white. Red indicates CKP> comparison group and blue indicates CKP<comparison group. **Abbreviations:** CKP=Chronic Knee Pain; CBP=Chronic Back Pain; CHP=Chronic Hip Pain; CNSP =Chronic Neck/Shoulder Pain; l=left; r=right; MF=multifidus; ES=erector spinae; PM=psoas major; QL=quadratus lumborum; GMin=gluteus minimus; GMed=gluteus medius; GMax=gluteus maximus; TFL=tensor fasciae latae; VL=vastus lateralis; VI=vastus intermedius; VM=vastus medialis; RF=rectus femoris; Sar=sartorius; Gra=gracilis; SM=semimembranosus; ST=semitendinosus; BFLH=biceps femoris long head; BFSH=biceps femoris short head; AdM=adductor magnus; AdL=adductor longus; AdB=adductor brevis.

### S4. Pairwise Group Differences between Single-Site Chronic vs Acute Pain

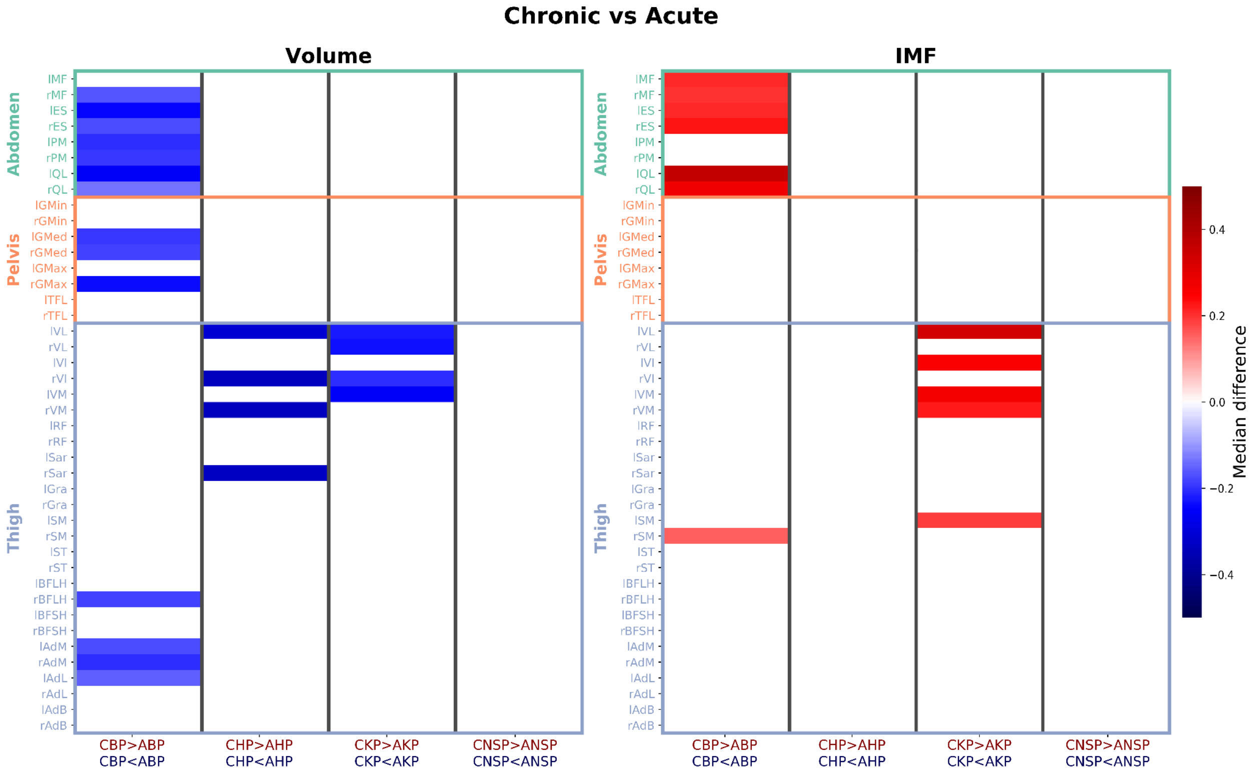


**Figure S5. Pairwise group differences in muscle Z-scores.** The heatmap shows median differences in normative Z-scores comparing participants with chronic vs acute pain groups. Rows correspond to 42 bilateral muscle ROIs, grouped by region. Columns show the pairwise comparisons against the CKP group (CBP vs ABP, CHP vs AHP, CKP vs AKP, CNSP vs ANSP). Cell color denotes the median Z-score difference for each comparison, restricted to results surviving Benjamini–Hochberg FDR correction applied within each pairwise comparison across 42 muscles ($q < .05$); non-significant cells are shown in white. Red indicates chronic > acute and blue indicates acute < chronic. **Abbreviations:** CKP=Chronic Knee Pain; CBP=Chronic Back Pain; CHP=Chronic Hip Pain; CNSP =Chronic Neck/Shoulder Pain; AKP=Acute Knee Pain; ABP=Acute Back Pain; AHP=Acute Hip Pain; ANSP =Acute Neck/Shoulder Pain; l=left; r=right; MF=multifidus; ES=erector spinae; PM=psoas major; QL=quadratus lumborum; GMin=gluteus minimus; GMed=gluteus medius; GMax=gluteus maximus; TFL=tensor fasciae latae; VL=vastus lateralis; VI=vastus intermedius; VM=vastus medialis; RF=rectus femoris; Sar=sartorius; Gra=gracilis; SM=semimembranosus; ST=semitendinosus; BFLH=biceps femoris long head; BFSH=biceps femoris short head; AdM=adductor magnus; AdL=adductor longus; AdB=adductor brevis.

### S5. Patterns of Muscle Health with Increasing Number of CPS

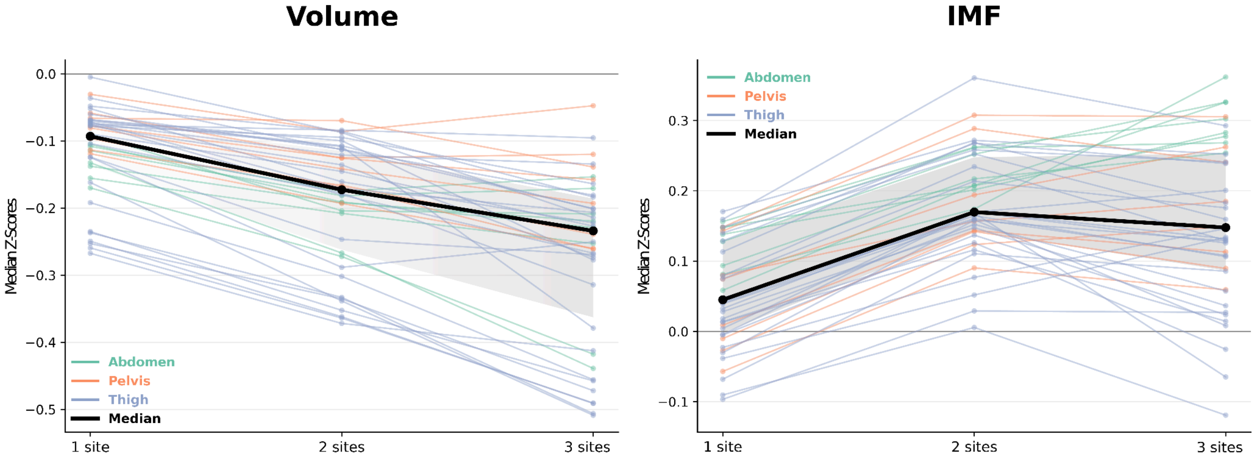


**Figure S6. Z-score trends of muscle health with increasing number of chronic musculoskeletal pain sites (CPS).** Line plots show median Z-scores for muscle volume and intramuscular fat (IMF) across the 1, 2, and 3 CPS, with thin colored lines showing individual muscles by anatomical region (abdomen, pelvis, thigh), the thick black line shows the median across all 42 muscles, and the gray shaded band indicates the interquartile range across muscle-wise median Z-scores. The horizontal reference line denotes Z=0.

**Table S3. Significant deviations from the norm in muscle volume and IMF across multi-site chronic pain groups.** The table shows all significant results derived from the normative modeling analysis for chronic pain groups. For each group and outcome (volume, IMF), the muscle abbreviation, median deviation, 95% confidence interval, and Bonferroni-corrected p-value are listed. Only results surviving Bonferroni-corrected significance threshold (p <.05) are shown. **Abbreviations:** l=left; r=right. MF=multifidus; ES=erector spinae; PM=psoas major; QL=quadratus lumborum; GMin=gluteus minimus; GMed=gluteus medius; GMax=gluteus maximus; TFL=tensor fasciae latae; VL=vastus lateralis; VI=vastus intermedius; VM=vastus medialis; RF=rectus femoris; Sar=sartorius; Gra=gracilis; SM=semimembranosus; ST=semitendinosus; BFLH=biceps femoris long head; BFSH=biceps femoris short head; AdM=adductor magnus; AdL=adductor longus; AdB=adductor brevis.

| Multi-Site Chronic Musculoskeletal Pain | | | | | |
|---|---|---|---|---|---|
| **Group** | **Outcome** | **Muscle** | **Median Deviation** | **95% CI** | **p (corr.)** |
| 1 CPS | Volume | lMF | -0.11 | -0.14 – -0.07 | <.001 |
| 1 CPS | Volume | rMF | -0.11 | -0.15 – -0.08 | <.001 |
| 1 CPS | Volume | lES | -0.14 | -0.18 – -0.11 | <.001 |
| 1 CPS | Volume | rES | -0.11 | -0.15 – -0.07 | <.001 |
| 1 CPS | Volume | lPM | -0.13 | -0.17 – -0.09 | <.001 |
| 1 CPS | Volume | rPM | -0.17 | -0.20 – -0.13 | <.001 |
| 1 CPS | Volume | lQL | -0.16 | -0.19 – -0.11 | <.001 |

| | | | | | |
|---|---|---|---|---|---|
| 1 CPS | Volume | rQL | -0.09 | -0.13 – -0.06 | <.001 |
| 1 CPS | Volume | lGMin | -0.10 | -0.14 – -0.06 | <.001 |
| 1 CPS | Volume | rGMin | -0.08 | -0.12 – -0.04 | <.001 |
| 1 CPS | Volume | lGMed | -0.11 | -0.15 – -0.06 | <.001 |
| 1 CPS | Volume | rGMed | -0.12 | -0.15 – -0.09 | <.001 |
| 1 CPS | Volume | rGMax | -0.08 | -0.11 – -0.05 | <.001 |
| 1 CPS | Volume | lVL | -0.24 | -0.27 – -0.20 | <.001 |
| 1 CPS | Volume | rVL | -0.26 | -0.29 – -0.22 | <.001 |
| 1 CPS | Volume | lVI | -0.24 | -0.27 – -0.20 | <.001 |
| 1 CPS | Volume | rVI | -0.25 | -0.29 – -0.22 | <.001 |
| 1 CPS | Volume | lVM | -0.25 | -0.29 – -0.22 | <.001 |
| 1 CPS | Volume | rVM | -0.27 | -0.31 – -0.23 | <.001 |
| 1 CPS | Volume | lRF | -0.19 | -0.22 – -0.16 | <.001 |
| 1 CPS | Volume | rRF | -0.16 | -0.20 – -0.12 | <.001 |
| 1 CPS | Volume | rST | -0.08 | -0.11 – -0.05 | <.001 |
| 1 CPS | Volume | lAdM | -0.08 | -0.12 – -0.05 | <.001 |
| 1 CPS | Volume | rAdM | -0.10 | -0.14 – -0.06 | <.001 |
| 1 CPS | Volume | lAdL | -0.12 | -0.16 – -0.09 | <.001 |
| 1 CPS | Volume | rAdL | -0.12 | -0.16 – -0.09 | <.001 |
| 1 CPS | Volume | rBFLH | -0.09 | -0.13 – -0.05 | .002 |
| 1 CPS | Volume | rAdB | -0.07 | -0.12 – -0.03 | .005 |
| 1 CPS | Volume | rGra | -0.08 | -0.11 – -0.04 | .008 |
| 1 CPS | Volume | lAdB | -0.07 | -0.11 – -0.03 | .008 |
| 1 CPS | Volume | rSM | -0.07 | -0.11 – -0.03 | .009 |
| 1 CPS | Volume | rSar | -0.07 | -0.11 – -0.03 | .013 |
| 1 CPS | Volume | lSM | -0.07 | -0.10 – -0.03 | .020 |
| 1 CPS | Volume | lGMax | -0.07 | -0.11 – -0.03 | .024 |
| 1 CPS | Volume | lSar | -0.07 | -0.11 – -0.03 | .034 |
| 1 CPS | IMF | IMF | 0.16 | 0.11 – 0.20 | <.001 |

| | | | | | |
|---|---|---|---|---|---|
| 1 CPS | lMF | rMF | 0.15 | 0.11 – 0.19 | <.001 |
| 1 CPS | lMF | lES | 0.14 | 0.10 – 0.18 | <.001 |
| 1 CPS | lMF | rES | 0.15 | 0.10 – 0.20 | <.001 |
| 1 CPS | lMF | lQL | 0.09 | 0.05 – 0.14 | <.001 |
| 1 CPS | lMF | rQL | 0.13 | 0.09 – 0.17 | <.001 |
| 1 CPS | lMF | lGMed | 0.14 | 0.10 – 0.18 | <.001 |
| 1 CPS | lMF | rGMed | 0.15 | 0.10 – 0.19 | <.001 |
| 1 CPS | lMF | lVL | 0.15 | 0.10 – 0.19 | <.001 |
| 1 CPS | lMF | rVL | 0.13 | 0.08 – 0.17 | <.001 |
| 1 CPS | lMF | lVI | 0.11 | 0.08 – 0.16 | <.001 |
| 1 CPS | lMF | rVI | 0.14 | 0.09 – 0.18 | <.001 |
| 1 CPS | lMF | lVM | 0.16 | 0.12 – 0.21 | <.001 |
| 1 CPS | lMF | rVM | 0.17 | 0.13 – 0.22 | <.001 |
| 1 CPS | lMF | rGra | -0.09 | -0.13 – -0.05 | .003 |
| 1 CPS | lMF | rAdL | 0.08 | 0.04 – 0.12 | .003 |
| 1 CPS | lMF | lGra | -0.10 | -0.13 – -0.05 | .005 |
| 1 CPS | lMF | rGMax | 0.08 | 0.05 – 0.12 | .008 |
| 1 CPS | lMF | lPM | 0.08 | 0.04 – 0.13 | .010 |
| 1 CPS | lMF | lAdL | 0.07 | 0.03 – 0.11 | .018 |
| 1 CPS | lMF | lGMax | 0.07 | 0.03 – 0.13 | .029 |
| 2 CPS | Volume | lMF | -0.17 | -0.25 – -0.09 | <.001 |
| 2 CPS | Volume | rMF | -0.18 | -0.26 – -0.11 | <.001 |
| 2 CPS | Volume | lES | -0.19 | -0.25 – -0.13 | <.001 |
| 2 CPS | Volume | rES | -0.19 | -0.24 – -0.08 | <.001 |
| 2 CPS | Volume | lPM | -0.27 | -0.32 – -0.20 | <.001 |
| 2 CPS | Volume | rPM | -0.27 | -0.34 – -0.21 | <.001 |
| 2 CPS | Volume | lQL | -0.21 | -0.28 – -0.13 | <.001 |
| 2 CPS | Volume | rQL | -0.20 | -0.28 – -0.13 | <.001 |
| 2 CPS | Volume | lGMed | -0.17 | -0.24 – -0.11 | <.001 |

| | | | | | |
|---|---|---|---|---|---|
| 2 CPS | Volume | rGMed | -0.19 | -0.27 – -0.12 | <.001 |
| 2 CPS | Volume | lVL | -0.35 | -0.43 – -0.27 | <.001 |
| 2 CPS | Volume | rVL | -0.36 | -0.44 – -0.27 | <.001 |
| 2 CPS | Volume | lVI | -0.33 | -0.41 – -0.26 | <.001 |
| 2 CPS | Volume | rVI | -0.34 | -0.41 – -0.27 | <.001 |
| 2 CPS | Volume | lVM | -0.36 | -0.43 – -0.30 | <.001 |
| 2 CPS | Volume | rVM | -0.37 | -0.44 – -0.28 | <.001 |
| 2 CPS | Volume | lRF | -0.30 | -0.37 – -0.25 | <.001 |
| 2 CPS | Volume | rRF | -0.34 | -0.40 – -0.27 | <.001 |
| 2 CPS | Volume | lBFLH | -0.16 | -0.22 – -0.08 | <.001 |
| 2 CPS | Volume | rAdM | -0.18 | -0.26 – -0.11 | <.001 |
| 2 CPS | Volume | lAdL | -0.25 | -0.30 – -0.18 | <.001 |
| 2 CPS | Volume | rAdL | -0.29 | -0.38 – -0.21 | <.001 |
| 2 CPS | Volume | lAdB | -0.17 | -0.26 – -0.10 | <.001 |
| 2 CPS | Volume | rAdB | -0.18 | -0.26 – -0.11 | <.001 |
| 2 CPS | Volume | lAdM | -0.14 | -0.19 – -0.06 | .002 |
| 2 CPS | Volume | lGMin | -0.17 | -0.24 – -0.07 | .005 |
| 2 CPS | Volume | rGMin | -0.14 | -0.19 – -0.07 | .005 |
| 2 CPS | Volume | rBFLH | -0.15 | -0.22 – -0.07 | .005 |
| 2 CPS | Volume | rSM | -0.12 | -0.21 – -0.06 | .011 |
| 2 CPS | Volume | rSar | -0.11 | -0.17 – -0.04 | .021 |
| 2 CPS | Volume | lSM | -0.11 | -0.20 – -0.04 | .044 |
| 2 CPS | IMF | lMF | 0.26 | 0.16 – 0.34 | <.001 |
| 2 CPS | IMF | rMF | 0.26 | 0.18 – 0.35 | <.001 |
| 2 CPS | IMF | lES | 0.20 | 0.12 – 0.30 | <.001 |
| 2 CPS | IMF | rES | 0.21 | 0.14 – 0.32 | <.001 |
| 2 CPS | IMF | lPM | 0.17 | 0.10 – 0.29 | <.001 |
| 2 CPS | IMF | rPM | 0.21 | 0.12 – 0.32 | <.001 |
| 2 CPS | IMF | lQL | 0.22 | 0.12 – 0.30 | <.001 |

| | | | | | |
|---|---|---|---|---|---|
| 2 CPS | IMF | rQL | 0.25 | 0.17 – 0.35 | <.001 |
| 2 CPS | IMF | lGMed | 0.29 | 0.19 – 0.40 | <.001 |
| 2 CPS | IMF | rGMed | 0.31 | 0.21 – 0.40 | <.001 |
| 2 CPS | IMF | lGMax | 0.19 | 0.12 – 0.27 | <.001 |
| 2 CPS | IMF | lVL | 0.26 | 0.18 – 0.36 | <.001 |
| 2 CPS | IMF | rVL | 0.27 | 0.19 – 0.38 | <.001 |
| 2 CPS | IMF | lVI | 0.27 | 0.15 – 0.36 | <.001 |
| 2 CPS | IMF | rVI | 0.25 | 0.17 – 0.35 | <.001 |
| 2 CPS | IMF | lVM | 0.36 | 0.27 – 0.46 | <.001 |
| 2 CPS | IMF | rVM | 0.27 | 0.20 – 0.38 | <.001 |
| 2 CPS | IMF | rBFLH | 0.17 | 0.09 – 0.24 | <.001 |
| 2 CPS | IMF | lAdM | 0.17 | 0.09 – 0.26 | <.001 |
| 2 CPS | IMF | rAdM | 0.16 | 0.09 – 0.24 | <.001 |
| 2 CPS | IMF | lAdL | 0.21 | 0.14 – 0.31 | <.001 |
| 2 CPS | IMF | rAdL | 0.23 | 0.15 – 0.29 | <.001 |
| 2 CPS | IMF | rAdB | 0.17 | 0.08 – 0.26 | <.001 |
| 2 CPS | IMF | lSM | 0.17 | 0.08 – 0.25 | .002 |
| 2 CPS | IMF | rST | 0.15 | 0.06 – 0.21 | .002 |
| 2 CPS | IMF | rGMax | 0.16 | 0.07 – 0.25 | .003 |
| 2 CPS | IMF | lRF | 0.16 | 0.08 – 0.26 | .003 |
| 2 CPS | IMF | lBFLH | 0.15 | 0.07 – 0.21 | .003 |
| 2 CPS | IMF | lAdB | 0.14 | 0.07 – 0.24 | .003 |
| 2 CPS | IMF | rSM | 0.13 | 0.05 – 0.20 | .005 |
| 2 CPS | IMF | rGMin | 0.14 | 0.05 – 0.22 | .008 |
| 2 CPS | IMF | lGMin | 0.14 | 0.07 – 0.22 | .008 |
| 2 CPS | IMF | rRF | 0.16 | 0.06 – 0.24 | .013 |
| 2 CPS | IMF | rBFSH | 0.16 | 0.06 – 0.25 | .022 |
| 3 CPS | Volume | lPM | -0.44 | -0.59 – -0.29 | <.001 |
| 3 CPS | Volume | rPM | -0.42 | -0.55 – -0.28 | <.001 |

| | | | | | |
|---|---|---|---|---|---|
| 3 CPS | Volume | lVL | -0.46 | -0.60 – -0.33 | <.001 |
| 3 CPS | Volume | rVL | -0.49 | -0.62 – -0.35 | <.001 |
| 3 CPS | Volume | lVI | -0.51 | -0.65 – -0.38 | <.001 |
| 3 CPS | Volume | rVI | -0.47 | -0.65 – -0.28 | <.001 |
| 3 CPS | Volume | lVM | -0.49 | -0.59 – -0.37 | <.001 |
| 3 CPS | Volume | rVM | -0.41 | -0.59 – -0.29 | <.001 |
| 3 CPS | Volume | lRF | -0.46 | -0.60 – -0.32 | <.001 |
| 3 CPS | Volume | rRF | -0.51 | -0.64 – -0.34 | <.001 |
| 3 CPS | Volume | lSar | -0.28 | -0.38 – -0.13 | <.001 |
| 3 CPS | Volume | rAdL | -0.25 | -0.43 – -0.14 | <.001 |
| 3 CPS | Volume | lQL | -0.25 | -0.39 – -0.10 | .002 |
| 3 CPS | Volume | rSar | -0.38 | -0.53 – -0.19 | .002 |
| 3 CPS | Volume | lAdL | -0.27 | -0.40 – -0.15 | .002 |
| 3 CPS | Volume | rAdM | -0.31 | -0.48 – -0.12 | .006 |
| 3 CPS | Volume | lES | -0.22 | -0.34 – -0.11 | .007 |
| 3 CPS | Volume | lGMin | -0.26 | -0.44 – -0.13 | .008 |
| 3 CPS | Volume | lAdM | -0.27 | -0.39 – -0.10 | .008 |
| 3 CPS | Volume | lGMed | -0.24 | -0.39 – -0.07 | .029 |
| 3 CPS | Volume | rAdB | -0.27 | -0.37 – -0.08 | .030 |
| 3 CPS | Volume | lBFSH | -0.21 | -0.36 – -0.09 | .041 |
| 3 CPS | IMF | lPM | 0.36 | 0.21 – 0.53 | <.001 |
| 3 CPS | IMF | rPM | 0.33 | 0.17 – 0.47 | <.001 |
| 3 CPS | IMF | rQL | 0.33 | 0.22 – 0.47 | <.001 |
| 3 CPS | IMF | lVM | 0.29 | 0.11 – 0.48 | <.001 |
| 3 CPS | IMF | rGMed | 0.31 | 0.13 – 0.46 | .002 |
| 3 CPS | IMF | lMF | 0.30 | 0.14 – 0.44 | .003 |
| 3 CPS | IMF | rVM | 0.25 | 0.10 – 0.38 | .003 |
| 3 CPS | IMF | rMF | 0.27 | 0.14 – 0.43 | .005 |
| 3 CPS | IMF | rES | 0.28 | 0.11 – 0.41 | .007 |

| | | | | | |
|---|---|---|---|---|---|
| 3 CPS | IMF | lES | 0.28 | 0.11 – 0.45 | .008 |
| 3 CPS | IMF | rVL | 0.24 | 0.10 – 0.36 | .015 |

### S6. Predictive Value of Muscle Health Patterns in Single-Site Chronic and Acute Pain

To test the predictive value of normative markers, we trained a logistic regression model using muscle volume and IMF Z-scores as features to classify between the single-site chronic pain groups (i.e., three-class classification: back, hip, or knee). Model performance was evaluated using a nested cross-validation framework. The outer loop included a 10-fold stratified shuffle split, with 80% of the data used for training and 20% reserved for testing in each split while preserving class proportions. The inner loop was a fivefold stratified shuffle split of the training data (80%) from the outer loop, where hyperparameter optimization was performed. The optimized hyperparameters were selected using a Bayesian optimization search framework with a Tree-structured Parzen Estimator sampler. Feature standardization was implemented within the training set for each inner fold to prevent information leakage. The best-performing model was retrained on the full training set and evaluated on the held-out test data from the outer split, using macro-F1 and balanced accuracy. The overall performance estimates were summarized across outer splits. The statistical significance of the model performance was evaluated using permutation testing, in which a null distribution was generated by shuffling the data over 100,000 iterations. Only the outcome variable (y) was shuffled in permutation testing to break the feature-outcome association to generate the true null distribution. The feature matrix (X) is not shuffled to preserve the biologically meaningful relationships between muscle volume, IMF, and their spatial organization. Disrupting these underlying patterns would make the test less representative of the true data structure. This framework was also used to classify the single-site acute pain groups.

#### Ablation Analysis

Feature importance was assessed using ablation-based analyses. For single-feature ablation, values of each feature were replaced with zero in the training data, and the training process was repeated. Model performance after feature perturbation was compared with that of the full model, and the resulting decrease in predictive accuracy was used to estimate feature importance. This procedure was repeated for all features, yielding a computationally efficient approximation of how perturbations to individual predictors affect model performance.
